# Genetic liability to addiction underlies comorbid bipolar and substance use disorders

**DOI:** 10.64898/2026.02.04.26345483

**Authors:** Lars A. R. Ystaas, Pravesh Parekh, Nadine Parker, Ibrahim Akkouh, Viktoria Birkenæs, Ida E. Sønderby, Elise Koch, Espen Hagen, Oleksandr Frei, Alexey Shadrin, Ole A. Andreassen, Kevin S. O’Connell

## Abstract

**Background:** Bipolar disorder (BIP) frequently co-occurs with heightened substance use (SU) and substance use disorders (SUDs). Although the strong co-occurrence of these heritable traits points to shared genetic susceptibility, the extent to which there are differences in how SU and SUD overlap with BIP genetic architecture remains unclear.

**Methods:** We quantified the polygenic overlap between BIP and SUDs (alcohol, cannabis, opioid, and tobacco), and BIP and SU traits (drinks per week, lifetime cannabis use, prescription_opioid use, and smoking initiation) using GWAS summary statistics and trivariate MiXeR. We then isolated the general and unique genetic contributions of SUD and SU using GWAS-by-subtraction via GenomicSEM. Next, we tested associations between polygenic risk scores (PRSs) derived from these latent factors and diagnostic and behavioral outcomes in the Norwegian Mother, Father and Child Cohort Study. Finally, we applied GSA-MiXeR to explore pleiotropic pathway enrichment shared between the latent factors and BIP.

**Results:** We found extensive polygenic overlap between traits, with SUDs being more genetically correlated with BIP than SU traits. The unique SUD factor correlated positively with psychiatric disorders, whereas unique SU correlated negatively. PRSs for BIP, shared SUD/SU, and unique SUD were significantly associated with BIP, SUD, and comorbid SUD-BIP; PRS for unique SU was only associated with self-reported lifetime SU. GSA-MiXeR revealed richer gene-set enrichment for SUD/BIP than SU/BIP implicating dopamine signaling and interneuron function.

**Conclusion:** By dissecting the genetic liability to SUD and SU and investigating their relationship with BIP we find a genetic link driven by substance dependence but not substance use more broadly.

## Introduction

Bipolar disorder (BIP) is a severe mental disorder that can lead to a marked loss in quality of life, increased mortality and an array of negative health outcomes(1). It is highly heritable, with estimates ranging from 58-85% in family(2) and twin studies(3). The development and progression of BIP is heterogeneous, with variation in clinical characteristics and functional outcomes(4). BIP is frequently comorbid with a range of psychiatric and somatic conditions, of which substance use disorders (SUDs) are among the most common(5). When present, these disorders are associated with worse outcomes like increased symptom severity, more frequent affective episodes, poorer treatment response, and increased mortality(6,7). The most prevalent comorbid SUDs with BIP are alcohol use disorder (AUD, 42%), cannabis use disorder (CUD, 20%) and other illicit drug use disorder (17%)(8).

The high level of co-morbidity between BIP and SUD suggests that these disorders are etiologically linked. Similar to BIP, SUDs have high heritability, estimated between 40-72%(3), suggesting that a part of this etiological link is likely mediated through genetics.. Indeed, genomic studies of AUD, CUD, opioid (OUD), and tobacco (TUD) use disorders have found significant positive genetic correlations with BIP ranging from 0.08 to 0.30(9–12). The fact that these phenotypic and genetic associations are not restricted to any one particular substance, strongly suggests that there are common mechanisms driving the co-occurrence. Substantial effort has been made in recent years to explore the shared genetic risk among substance use (SU)-related behaviors. In genomic analyses, both pooling SUDs and externalizing behaviors, a general categorization of phenotypes related to under-controlled or impulsive action, has resulted in new insight(13–15). Of note, a phenome-wide association study (PheWAS) using a common SUD genetic addiction factor constructed using GenomicSEM, found that, apart from other SUDs, the strongest associations were with BIP(15).

While clinical characteristics of SUDs are often shared across substances, there is evidence that lifetime SU, or frequency measures of SU, show divergent genetic risk profiles compared to clinical SUDs(16–18). To take alcohol as an example, genetic liability to higher average number of drinks per week (DrnkWk) and alcohol use disorders (AUD) both increase the risk of having symptoms of AUD; however the effects are mediated through different motives: increased sociability and pleasant feelings for DrnkWk, and coping motives for AUD(19). Another example is how lifetime cannabis use (LCU) and CUD are respectively positively and negatively correlated with educational attainment(20,21). Recent work also highlights differential genetic links between substance use (SU) and substance-use disorder (SUD) within impulsivity. Sensation seeking and positive urgency - the tendency to act rashly when experiencing intense positive emotions - are more strongly tied to SU, whereas delay discounting - the preference for smaller, immediate rewards over larger, delayed ones - and negative urgency. - rash action driven by negative affect - are associated with SUD (22).

While accumulating genetic evidence thus supports the emerging concept that SU traits and SUDs are not a continuum, their disjoint genetic relationship with BIP is mainly unknown, as few studies have examined this directly. Because the genetic liabilities for SUDs and more general SU overlap extensively, we applied an analytical approach that separates the shared and unique, disorder-specific components in order to pinpoint relevant endophenotypes and clarify mechanisms driving BIP-substance comorbidity.

## Methods

### GWAS summary statistics

We used the most recent GWAS summary statistics with European ancestry for BIP, SUDs, and SU traits. The SUD summary statistics were for AUD(23), CUD(21), OUD(24), and TUD(12). The SU summary statistics were for number of drinks per week (DrnkWk)(25), lifetime cannabis use (LCU)(20,26), prescription opioid use (PrOU)(27), and smoking initiation (SmkInit)(25). We meta-analyzed LCU with MTAG as the outcomes were on different scales in the two input summary statistics. The summary statistics for BIP was the most recent BIP GWAS, excluding 23andMe self-report data, to focus on a more clinically severe phenotype(28), and were derived from a meta-analysis of cohorts excluding MoBa to avoid sample overlap. For more information about the GWAS summary statistics used in this study, see Supplementary Table 1.

### Bi- and Trivariate Gaussian Mixture Models (MiXeR)

We used MiXeR(29–31) to estimate polygenic overlaps between BIP and SU/SUD phenotypes. MiXeR estimates the proportion of shared trait-influencing variants between phenotypes, even if the effect directions are mixed. Bivariate and trivariate MiXeR are Gaussian mixture models that decompose genetic variance, understood as the distribution of SNP effects f3, into normally distributed components with polygenicity rr (the proportion of SNPs belonging to the component) and discoverability a^2^ (the average magnitude of the SNP effect size in the component). These components represent shared and trait-specific variants and null variants (variants with no effect on any of the analyzed traits).

### Genomic structural equation modeling (GenomicSEM)

We used genomic structural equation modeling (GenomicSEM)(32) to explore the genetic correlation between traits, and to generate SNP-level effects of underlying common genetic factors. We estimated genetic correlations with GenomicSEM’s implementation of LD-score regression(33). Genetic correlations presented in the text and supplementary tables are the output of the rgmodel function(34).

To focus on the unique genetic liabilities of SU and SUD respectively, we fit two separate two-level GWAS-by-subtraction models. GWAS-by-subtraction describes a model where the effect of one latent factor is removed from another, to create an orthogonalized residual factor(35). On the first level, we constructed common factors for SUD and SU traits. To model substance-specific effects, we allowed for correlations between residual variance in the indicator variables in the same substance class(36). On the second level, we constructed one factor (general SUD/SU) to capture genetic variance shared between SUD and SU, and one factor (unique SUD/SU) to capture unique non-shared variance for SU and SUD, respectively. For pathway enrichment analyses with GSA-MiXeR, we performed fixed effect meta-analyses between BIP and these GenomicSEM factors by setting the residual variance to zero and loadings to one. To evaluate model fit, we used comparative fit index (CFI) and standardized root mean residual (SRMR). We performed multivariable GWAS to estimate SNP-level effects of our latent factors to be used in downstream analyses, excluding SNPs that showed signs of heterogeneity across input traits. For more details, see supplement.

### Norwegian Mother, Father and Child Cohort Study (MoBa)

We utilized genotype data from the Norwegian Mother, Father and Child Cohort Study (MoBa)(37) to investigate the genetic and phenotypic relationships between BIP and SUD. MoBa is a population-based pregnancy cohort study where all pregnant mothers, their offspring and the fathers were eligible to join in the period 1999-2008. The women consented to participation in 41% of the pregnancies, for a total of approximately 114,500 children, 95,200 mothers and 75,200 fathers. For this work, consenting participants as of 25th April 2025, encompassed a final sample size of 112,475 children, 94,375 mothers and 74,295 fathers. We used the quality controlled genotyping data made available from the MoBaPsychGen pipeline v1(38), consisting of a final sample of 207,569 individuals who passed all QC steps. Details on phenotypes drawn from questionnaire and registry data are included in supplementary methods. MoBa is regulated by the Norwegian Health Registry Act. The current study was approved by The Regional Committees for Medical and Health Research Ethics (2016/1226/REK Sør-Øst C).

### Polygenic risk score analysis

To investigate the genetic disposition associated with comorbid BIP and SUD among MoBa participants, we generated Polygenic Risk Scores (PRS) from BIP GWAS summary statistics and the four GenomicSEM latent factors: General SUD, Unique SUD, General SU, Unique SU. We used the PRS-PCA method first published by Coombes et al to calculate the scores(39). We first calculated a PRS for each participant at 11 p-value thresholds ranging from 5e^−8^ to 1.0 using PRSice-2(40). Next, we extracted the first principal component from the correlation matrix of these 11 PRS’ and used this for downstream analyses.

We then evaluated the strength of the association between the PRS’ and diagnostic outcomes bipolar disorder (BIP, ICD-10 F31), SUD (any ICD-10 F1 chapter, excluding F1x.0 acute intoxications; or the ICPC-2 codes P15 chronic alcohol abuse, P17 tobacco abuse, P18 prescription medication abuse or P19 substance abuse), as well as F31/SUD comorbidity. Models were fit with population, BIP and SUD controls. Population controls were not screened for any characteristic. We used the following prevalence numbers to estimate liability scale pseudo-R^2^: 0.02 for BIP(4), 0.10 for SUD(41,42), and 0.50 for comorbid SUD in BIP(8). We also explored our genetic factors’ associations with self-reported substance use in adults (from the questionnaire the parents fill out in week 15 of the pregnancy) and in adolescents (from the questionnaire participants fill out as 14 year-olds). We finally looked at associations with BIP diagnostic subtypes: Individuals with lifetime diagnosis of F30/F31 hypomania, but no mania; and those with lifetime history of mania. These groups were chosen to be proxies for type 2 and type 1, respectively. We also looked at associations with treatment-resistant BIP, defined as lifetime history of BIP and either complex polypharmacy (concurrent use of ≥3 BIP drugs within a 180-day period) or lifetime use of clozapine. There is no consensus definition of treatment-resistance in BIP, but this definition conforms with recommendations in other literature (43,44). We fit logistic regression models using the function lrm(45). Our main focus in the discussion are the models including BIP and two latent factors from the same GWAS-by-subtraction model, but nested models were fit in order to do inference on parameters. Hypothesis testing of the PRS was done via likelihood ratio test, and Benjamini-Hochberg adjusted p-values were considered significant under a threshold of 0.05. All models were fit with covariates age, sex, 20 first genetic principal components and 6 imputation batches as covariates. Bootstrapped confidence intervals were acquired using the package boot(46) and 1000 bootstrap iterations. Percentile confidence intervals were used for pseudo- R^2^, and studentized confidence intervals for regression coefficients.

### Gene set analysis with GSA-MiXeR

We mapped our fixed effect meta-analyses between BIP and GenomicSEM multivariable GWAS factors to genes and gene-sets using GSA-MiXeR(47). GSA-MiXeR is a functional mapping tool that prioritizes smaller and more specific gene sets compared to other gene- and gene set mapping approaches. The GSA-MiXeR model extends the MiXeR framework described above by using gene-and functional annotations to reduce the number of parameters necessary to model the genetic variance. It estimates heritability enrichment in a particular annotation by comparing a full model with varying discoverability depending on gene, to a base model where all SNPs have the same variance parameter regardless of which gene region they inhabit. We used a Haplotype Reference Consortium (HRC) reference like in the original publication, as well as the same gene boundaries, gene sets and functional annotations. The boundaries of each gene were extended by 10 kb up- and down-stream. We considered a gene set implicated if it had an AIC>0 and enrichment > 1 + enrichment standard error.

## Results

### Genetic architecture of individual SUD and SU phenotypes

Genetic correlations varied between a low of 0.08 (TUD) to a high of 0.35 (LCU) (Supplementary Table 2), with SUDs generally being more correlated with BIP. All phenotypes showed considerable polygenicity as estimated by univariate MiXeR, ranging from 5.3k trait-influencing variants for OUD to 12.0k for TUD, with SU traits in general more polygenic than SUDs (Supplementary Table 3). Polygenic overlap estimated by bivariate and trivariate MiXeR was extensive, with only a minority of trait-influencing variants being unique to any one disorder (Figure 1, Supplementary Figure 1A, Supplementary Table 4 and 5). The relationship between SUD and SU phenotypes ranged from the large overlap in alcohol, where AUD trait-influencing variants largely were a subset of the DrnkWk variants, to opioids, where 53% of the total number of trait-influencing variants associated with PrOU were not shared with OUD, and 33% of those associated with OUD were not shared with PrOU.

**Figure 1.**
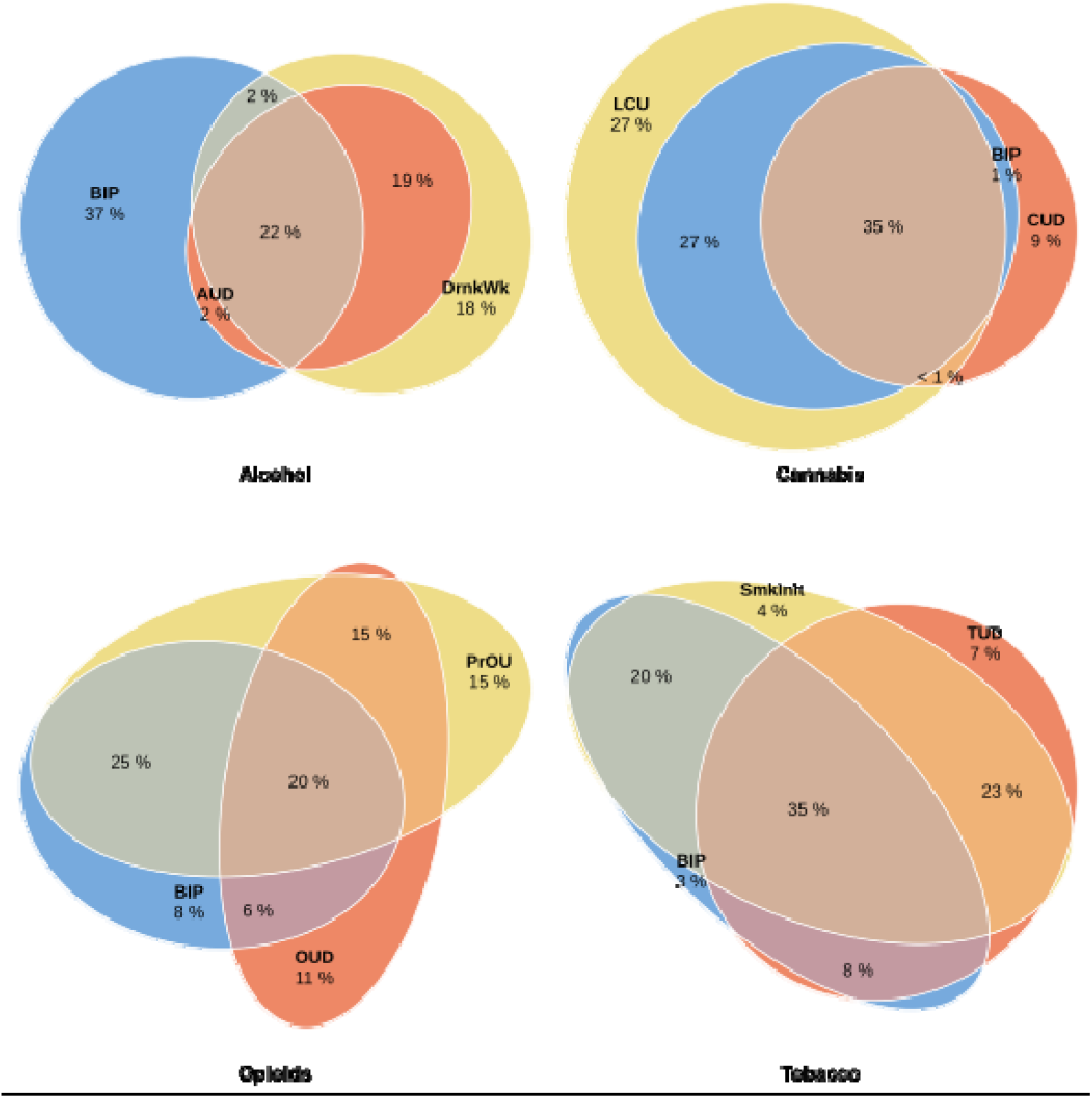
Genetic overlap between BIP, SUD and SU traits as estimated by Trivariate MiXeR. The percentages show the proportion of trait-influencing variants within each section of the Venn diagram relative to the sum of all trait-influencing variants. The size of the circles reflects the polygenicity of each trait. Abbreviations: BIP (bipolar disorder), AUD (alcohol use disorder), DrnkWk (Drinks per week), CUD (cannabis use disorder), LCU (lifetime cannabis use), OUD (opioid use disorder), PrOU (prescription opioid use), TUD (tobacco use disorder), SmkInit (smoking initiation).

### Genetic architecture of common genetic factors

To move beyond relationships between single phenotypes and explore the common underpinnings of the phenotypes in question, we fit two theory-driven GenomicSEM models to SUD and SU summary statistics (Figure 2A, Supplementary table 6). The two-level models estimated latent common factors for SUD and SU on the first level, and on the second level, a general factor loading on SUD and SU as well as a unique SUD/SU factor loading on the residual variance in either SUD or SU, respectively. Both models showed adequate fit (CFI=0.95, SRMR=0.08). As could be expected from the weaker correlations between SU traits, the SU factor showed signs of heterogeneity, with weaker factor loadings and more residual variance in the indicator variables. When analyzing the latent factors with MiXeR, the unique SU and SUD factors were less polygenic than their general counterparts (Figure 2C, Supplementary table 5). In all cases, there was substantial overlap with BIP.

**Figure 2.**
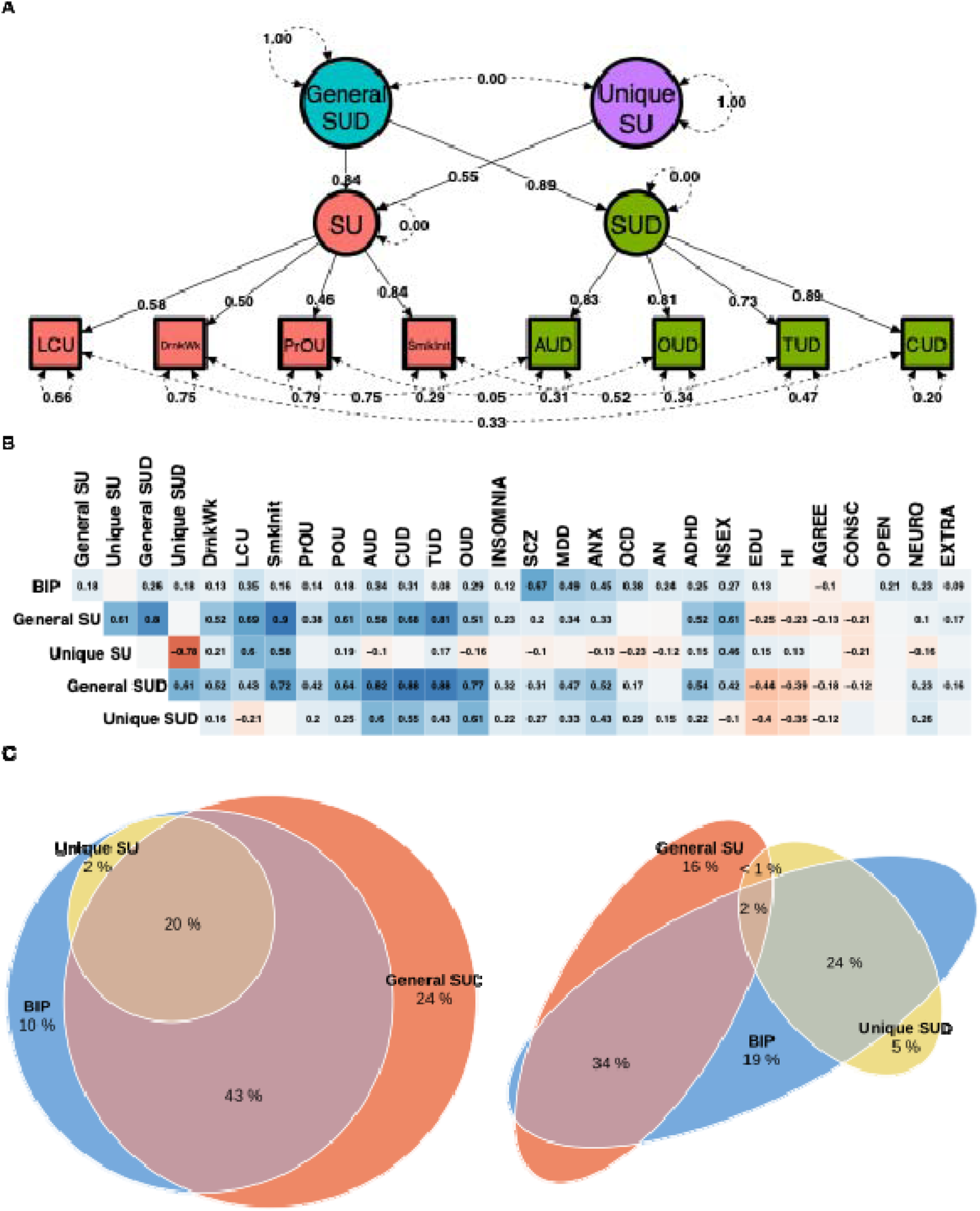
A: GenomicSEM modeling: 2-level GWAS-by-subtraction on SUD and SU phenotypes. General SUD/Unique SU is used as an example, with the reverse model being identical aside from general SU loading on both SU and SUD, and unique SUD loading only on SUD. On the first level, common factors for SUD and SU traits are constructed. On the second level, a general SUD factor captures all common variance in SUD/SU phenotypes, while a residual unique SU factor is defined to extract residual variance in SU. Residual variance in the indicator variables is allowed to correlate for each substance class. B: Genetic correlations between latent factors, BIP and other relevant psychiatric and behavioral traits. All cells with coefficients were statistically significantly different from zero at a p-value threshold of 0.05 after Benjamini-Hochberg correction. Phenotypes included: POU (problematic opioid use)(48), SCZ (schizophrenia)(49), MDD (Major depressive disorder)(50), ANX (anxiety disorder)(51), OCD (Obsessive-compulsive disorder)(52), AN (anorexia nervosa)(53), ADHD (attention deficit hyperactivity disorder)(54), NSEX (lifetime number of sexual partners)(55), EDU (educational attainment)(56), HI (household income)(57), AGREE/CONSC/OPEN/NEURO/EXTRA (personality traits agreeableness, conscientiousness, openness, neuroticism, extraversion)(58). C: MiXeR analysis of BIP and latent factors.

To understand what these latent factors represent, we first looked to genetic correlations with a selection of relevant psychiatric and behavioral traits (Figure 2B, Supplementary table 2). The unique SU factor showed only weak to absent correlations with SUDs, while being strongly linked to the specific use measures LCU, SmkInit and DrnkWk. It was further positively correlated with educational attainment and household income, while being negatively correlated with neuroticism, obsessive-compulsive disorder (OCD) and anorexia nervosa. In contrast, the unique SUD factor was strongly correlated with SUDs and showed weak to negative correlations with SU traits. Unique SUD displayed general positive correlations with psychiatric traits such as schizophrenia, insomnia, anxiety, MDD, and ADHD; as well as neuroticism; together with negative correlations for education, income and agreeableness. A distinctive feature of the unique SUD factor was its positive genetic correlation with both OCD and anorexia nervosa and its negative correlation with lifetime number of sexual partners (NSEX) – the positive correlation with the compulsive traits being stronger than the general SUD factor, and the negative correlation with the impulsivity-related trait NSEX having an opposing sign to that of all other latent factors.

The general factors showed largely similar correlation profiles with each other. They had broad positive correlations with both SU traits and SUDs, moderate to strong correlations with psychiatric disorders, positive correlations with extraversion, and negative correlations with education, income, agreeableness and conscientiousness.

### Validation of genetic associations in MoBa

We next aimed to validate our genetic factors by exploring polygenic associations in the MoBa cohort. For the main diagnostic outcomes BIP, SUD, and BIP/SUD comorbidity (Figure 3, Supplementary Figure 3A/4E, Supplementary table 7), we found that a higher BIP PRS was associated with an increased lifetime risk of both BIP and SUD diagnoses relative to population controls. BIP PRS also raised the likelihood of BIP/SUD comorbidity when compared with individuals who had only SUD, but it did not distinguish comorbid cases from those with only BIP. Unique SUD PRS was predictive of SUD against both population and BIP controls, although with a smaller effect in the latter case, consistent with it also being predictive of BIP against population controls. Unique SUD PRS was also associated with higher risk of BIP/SUD comorbidity against population and BIP controls, but not against SUD controls. General SU and SUD had similar patterns, but with generally larger effects, especially for general SUD. Unique SU was significantly associated with SUD against both population and bipolar controls, but the effect as measured by incremental R^2^ was marginal (Supplementary Figure 3A) .

**Figure 3.**
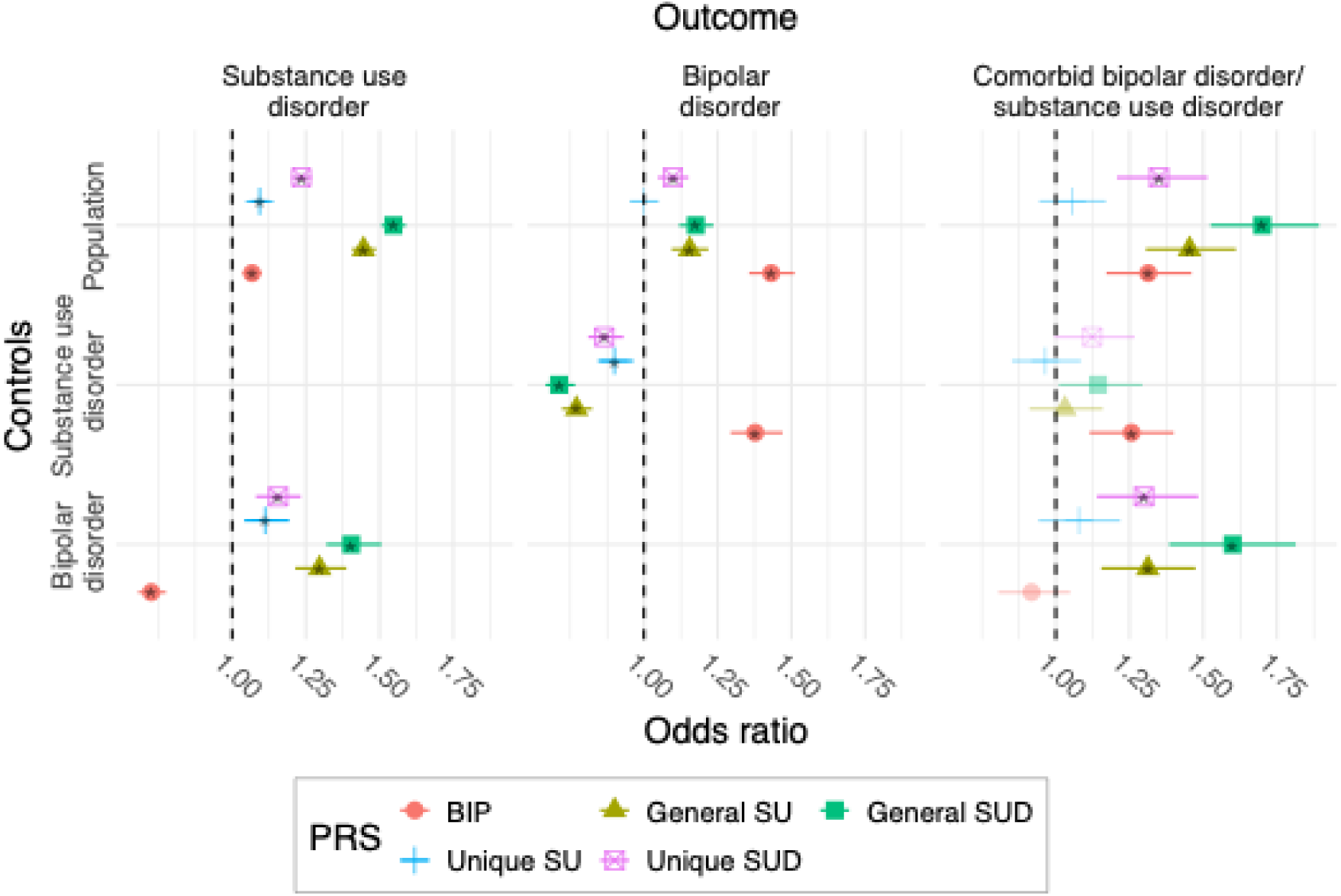
PRS associations with diagnosis in MoBa, against population, SUD or BIP controls. Error bars denote bootstrap confidence intervals. Odds ratio for 1 SD. increase in PRS value. Stars denote statistical significance. More information is provided in supplementary table 7.

We then analyzed SU as indexed by self-reported lifetime use in MoBa questionnaire data (Figure 4, Supplementary Figure 3C/3D/3G/3H, Supplementary table 7). The effect of BIP genetic risk was muted, and only had a significant effect on lifetime cannabis use in adults. In contrast, general SUD/SU and unique SU were associated with increased probability of lifetime use of all included substances in both adults and adolescents. Unique SUD was only significantly associated with tobacco use in the adult population, and even then showed marginal incremental R^2^ (Supplementary Figure 3H). The effects were largely similar across diagnostic groups.

**Figure 4.**
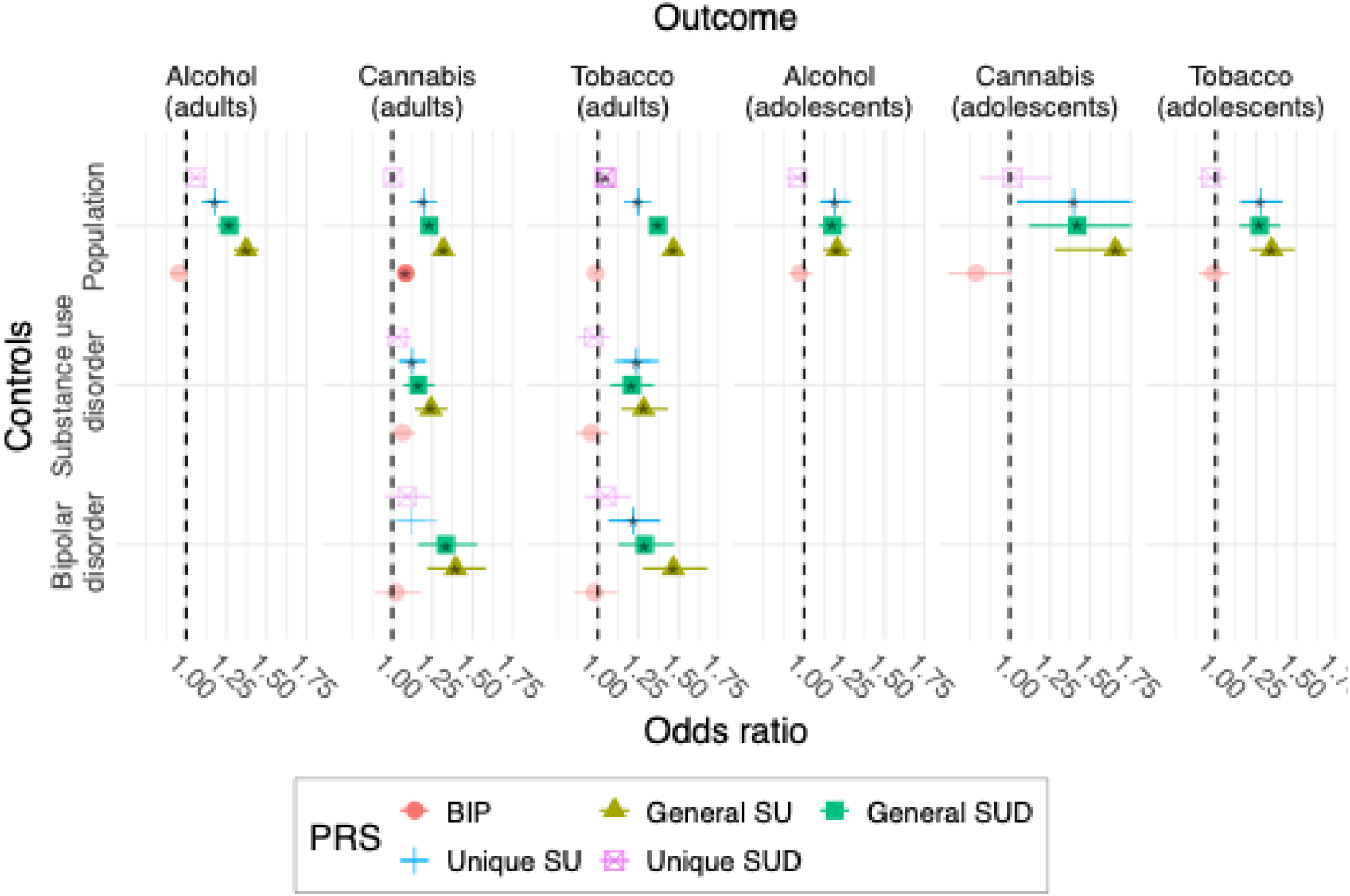
PRS associations with self-reported lifetime substance use in MoBa adults and adolescents within general population, BIP and SUD cases. Error bars denote bootstrap confidence intervals. Due to small numbers of individuals in the dataset without lifetime alcohol use and adolescents in diagnostic subgroups, models in subgroups for these phenotypes were not able to be estimated. Odds ratio for 1 SD. increase in PRS value. Stars denote statistical significance. More information is provided in supplementary table 7.

To explore the genetic loading in different subtypes of BIP, we fit models where the outcome was defined as lifetime diagnosis of mania, as lifetime diagnosis of hypomania but no mania, or as treatment-resistant bipolar (Figure 5, Supplementary Figure 3B/3F, Supplementary table 7). As expected, a higher BIP PRS was associated with increased risk of both mania and hypomania in population controls. A higher BIP genetic risk was also seen in the treatment-resistant bipolar group when compared to the population, but this was not the case compared to other BIP cases. The Unique SUD factor had a significant effect on increasing the risk of treatment-resistant bipolar in the bipolar group.

**Figure 5.**
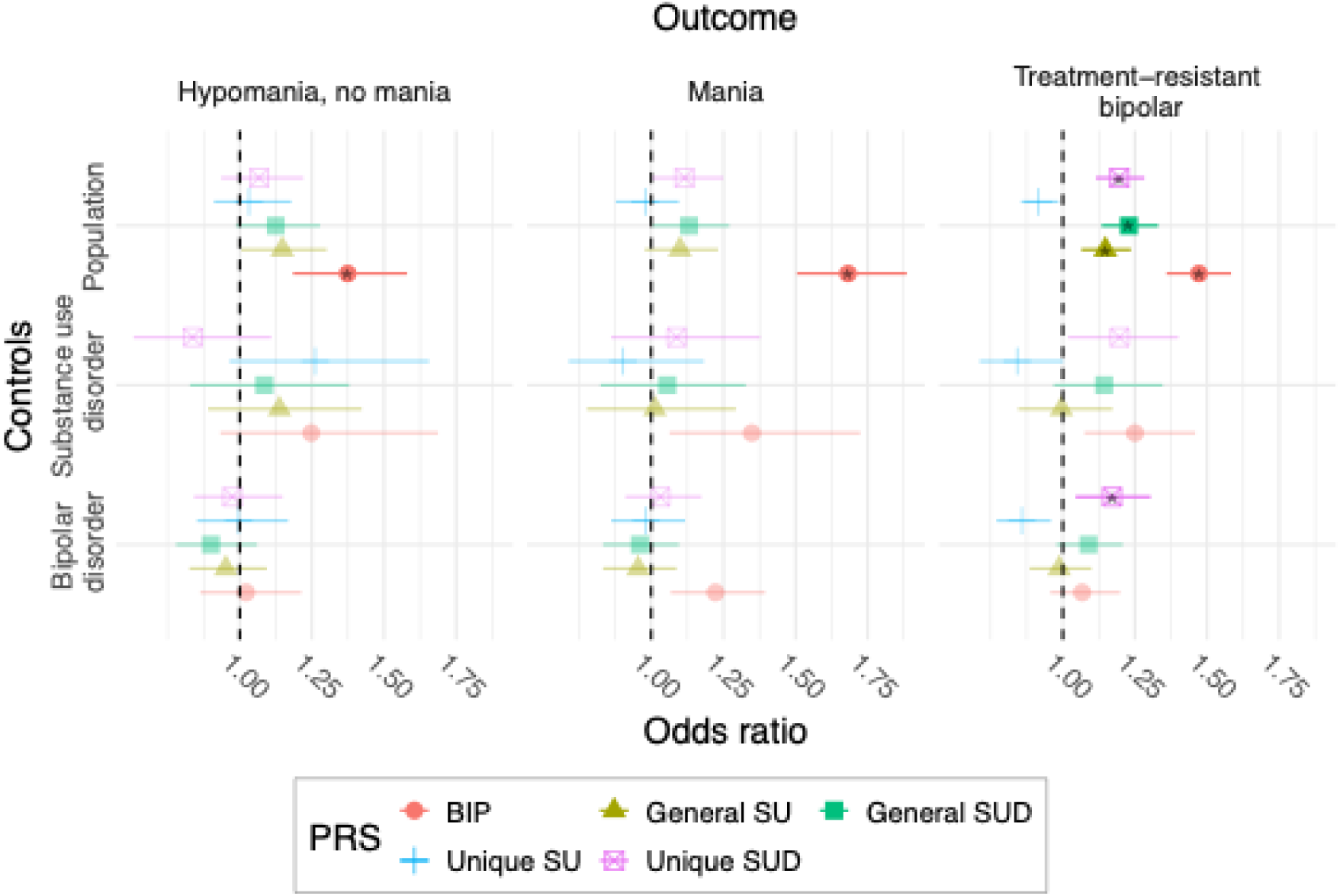
PRS associations with BIP diagnostic subtypes in MoBa participants. “Hypomania, no mania” refers to individuals with lifetime diagnosis of hypomania, but no mania diagnosis, while “Mania” refers to anyone with a lifetime diagnosis of mania. Error bars denote bootstrap confidence intervals. Odds ratio for 1 SD. increase in PRS value. Stars denote statistical significance. More information is provided in supplementary table 7.

### Gene set analysis with GSA-MiXeR and MAGMA

To further explore the biological correlates of the shared variance between BIP and our SUD/SU factors, we performed a fixed effects meta-analysis in GenomicSEM, followed by gene set analysis with GSA-MiXeR(Figure 6, Supplementary table 8). The meta-analyses including general SUD had the most implicated gene sets with 14, while the meta-analysis with unique SUD had the greatest mean fold enrichment. Seven implicated gene sets were shared between unique and general SUD, mostly related to dopamine signaling. The meta-analyses including general and unique SU implicated 8 and 2 enriched gene sets, respectively, none of which were implicated in the general and unique SUD analyses. The SU implicated gene sets also showed lower fold enrichment, and most were either unspecific (e.g. “Positive regulation of 3-UTR-mediated mRNA stabilization”) or related to neurodevelopment (e.g. “Commisural neuron axon guidance”).

**Figure 6.**
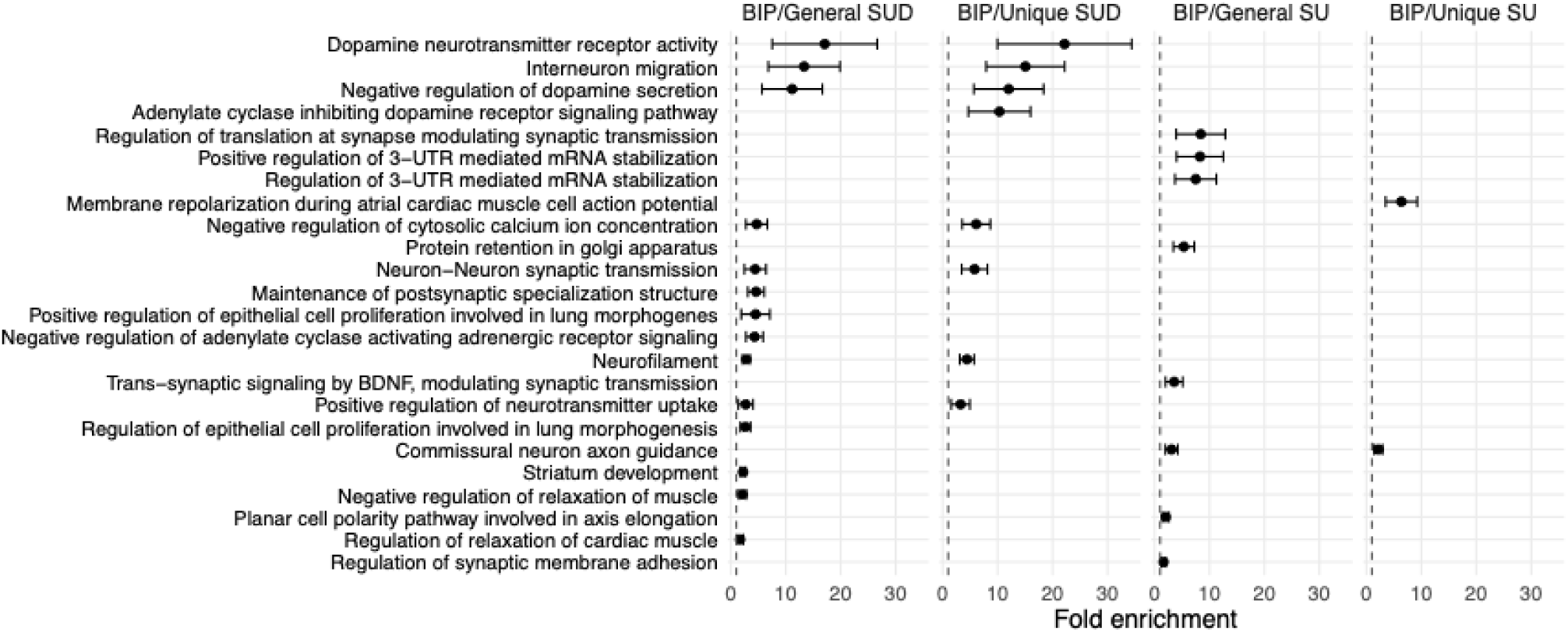
Gene set enrichment as determined by GSA-MiXeR. The dot markers show fold enrichment, while error bars show standard errors of the model estimates. Gene sets are sorted by mean enrichment across meta-analyses. Vertical dotted line denotes 1, i.e., no enrichment.

## Discussion

Our analyses of the common genetic underpinnings of BIP, SU, and SUD show a distinct pattern of different genetic architecture. When we generate latent factors using GWAS-by-subtraction to capture the shared variance between SUDs and SU, these are predictive of both SUD and self-reported SU in the MoBa cohort, while unique SUD/SU factors only predict clinical SUD or self-reported SU, respectively. Of these, the unique SUD but not SU factor is correlated with BIP and shows pathway enrichment when meta-analyzed with BIP. Finally, when we analyze subgroups of BIP, we find that the group with comorbid SUD is distinguished by elevated SUD and not BIP PRS, and that unique SUD has an association with treatment resistance that is not picked up by the general factors. Our analyses suggest three major conclusions: i) Even though there is a large degree of overlap and correlation between SU and SUD traits, there remains genetic variance that is specific to either group of traits. ii) BIP displays more genetic overlap with variance specific to SUD compared with SU. iii) Despite the close genetic relationship between BIP and SUD, comorbid SUD should not be considered as simply another symptom of BIP, but rather an important subgroup with overlapping but distinct genetic risk.

The unique SU factor appears to be “non-pathological” in its correlation with other phenotypes. Genetic correlations show low to negative correlations with SUDs, psychiatric disorders, neuroticism and insomnia; while there is weak, positive correlation with educational attainment and household income. Further supporting its non-pathological profile, its PRS is predictive of self-reported SU across a wide range of substance classes in adolescents and adults, while it is not associated with SUD. This is in line with previous literature which suggest that the relationship between SU behaviors and social and health outcomes is context-dependent(59). For instance, effects of early adolescent alcohol consumption on later adverse outcomes are modulated and confounded by culture, parental drinking behavior, impulsivity and other externalizing problem behaviors(60). A Norwegian longitudinal study found that the effect was age-dependent, with drunkenness in the early teens being predictive of lower education and income later in life, while drunkenness in the twenties and thirties having a positive association with these outcomes(61). The authors suggest that participation in normative SU behaviors might both be an indicator of social integration and function and have beneficial effects on later employment and educational opportunities through networking.

In contrast, our unique SUD factor is genetically correlated with SUDs, mental disorders and inversely correlated with education and income. It is particularly distinguished among our SUD/SU factors by having the highest genetic correlation with the compulsive disorders OCD and anorexia nervosa. The relationship between OCD and SUD is controversial. A meta-analysis in clinical samples found reduced prevalence rates of comorbid SUD with OCD(62), but a large-scale longitudinal population-based study in Sweden found increased risk of SUD for people with OCD, and estimated 55-68% of the covariance to be caused by genetic factors(63). OCD is also frequently comorbid with BIP (64), with obsessive-compulsive symptoms (OCS) restricted to, or worsening under, depressive episodes(65,66). The impulsivity subfactor negative urgency, the tendency to act rashly when experiencing extreme negative emotions, has been found to be associated with both SUD and OCS(67) and to distinguish SUD from SU liability in genomic analyses(22). The fact that our unique SUD/SU factors diverge in respect to compulsive traits, and also in their effect on our pragmatic definition of treatment-resistance, suggest that compulsivity, possibly via negative urgency, is a symptom dimension relevant to clinical outcomes in BIP. In this regard, the unique SUD factor might be indexing pleiotropic effects not limited to changes in SU behaviors, as the literature on pharmacological treatment of BIP reveals an ambiguous relationship with SU, with other comorbid mental disorders being more predictive of polypharmacy(68).

Functional enrichment analysis of shared pathways between BIP and the four factors show a clear difference in the level and specificity of enrichment, with SUD/BIP showing a higher level of enrichment more specific to the central nervous system. Dopamine signaling is a well-known component of SUD/SU behaviors, as well as BIP and psychotic disorders(4,69). Interneuron migration has also been implicated in BIP, schizophrenia and depression, with dysfunction of GABAergic interneurons in the cortex and hippocampus leading to disruption of the excitation-inhibition balance, which has negative consequences for cognitive function and a possible role in the other symptoms of the disorders(70–72). The majority of the gene sets enriched in the BIP/general SU meta-analysis are pathways related to basic cellular processes and neural development. The three most enriched in this analysis are all related to mRNA stabilization by RNA-binding proteins, and driven by the gene *ELAVL4*. RNA-binding proteins have an essential role in regulating gene expression, and the *ELAVL* gene family has established connections to behavioral abnormalities and neurological disease(73).

Our study has limitations. Although GWAS of clinical SUD with large sample size in the four substance classes we study have now been published, the same is not true for SU. Of particular note, an adequately powered GWAS of recreational opioid use was not available when performing the analyses. Medium- to long-term prescription opioid use (PrOU) is indicative of at least an effective and tolerable response to the drug, but clearly stands out from the other traits grouped under SU. This could introduce some heterogeneity into our construction of a SU factor and reduce power. The large discrepancies in power between the GWASs we incorporate in our SU latent factors also means that certain traits (i.e. SmkInit) have an outsized effect on the latent factor. Another limitation is that we restrict our analyses to European ethnicity, which reduces the generalizability of our findings. This is due to the lack of well-powered GWAS in non-European ethnicities for all the included phenotypes. Finally, the cultural and environmental variations in substance use globally require care in interpreting the implications of our results.

In conclusion, our findings delineate distinct genetic factors related to SU and SUD in both adolescence and adulthood. While we observe an overlap in heritable variance between SU and SUD, we demonstrate that the genetic risk associated with BIP does not correlate with SU-related variance that is separate from SUD. Instead, it appears that BIP genetic risk is more closely aligned with factors predisposing individuals to addiction and harmful substance use, rather than heightened experimentation or sensation seeking behaviors. Notably, individuals with comorbid BIP and SUD exhibit a higher relative genetic risk for our unique and general SUD factors compared to BIP, suggesting that for many in this group, their symptoms and behaviors may be more influenced by genetic predisposition to substance use rather than representing an exacerbation of BIP. These insights could have significant implications for understanding the complex interplay between genetic factors, substance use, and mood disorders.

## Data Availability

Publicly available GWAS summary statistics are available from the original authors or GWAS catalog. Data from MoBa used in this study are managed by the national health register holders in Norway (NIPH) and can be made available to researchers, provided approval from REC, compliance with the EU General Data Protection Regulation (GDPR), and approval from the data owners. The consent given by the participants does not open for storage of data on an individual level in repositories or journals. Researchers who want access to data sets for replication should apply through helsedata.no. Access to data sets requires approval from REC in Norway and an agreement with MoBa.

## Acknowledgements and funding information

The Norwegian Mother, Father and Child Cohort Study is supported by the Norwegian Ministry of Health and Care Services and the Ministry of Education and Research. We are grateful to all the participating families in Norway who take part in this on-going cohort study. We thank the Norwegian Institute of Public Health (NIPH) for generating high-quality genomic data. We also thank the NORMENT Centre for providing genotype data, funded by the Research Council of Norway (#223273), Southeast Norway Health Authorities, deCODE Genetics, the HARVEST collaboration and Stiftelsen Kristian Gerhard Jebsen. We further thank the Center for Diabetes Research, the University of Bergen for providing genotype data and performing quality control and imputation of the data funded by the ERC AdG project SELECTionPREDISPOSED, Stiftelsen Kristian Gerhard Jebsen, Trond Mohn Foundation, the Research Council of Norway, the Novo Nordisk Foundation, the University of Bergen, and the Western Norway Health Authorities. The current work was performed on Services for sensitive data (TSD), University of Oslo, Norway, with resources from UNINETT Sigma2 - the National Infrastructure for High-Performance Computing and Data Storage in Norway. A version of the manuscript has been presented in poster form at the World Congress of Psychiatric Genetics 2024 and International Society of Bipolar Disorder 2025. We gratefully acknowledge support from the European Union’s Horizon 2020 Research and Innovation Programme (RealMent; #964874, CoMorMent project; Grant #847776; Marie Skłodowska-Curie grant; #801133), the European Economic Area and Norway Grants (EEA-RO-NO-2018-0573), the National Institutes of Health (NIH 5R01MH124839-02, U24DA041123 and U24DA055330), the Research Council of Norway (#296030, #326813, #334920 and #324252), the South-Eastern Norway Regional Health Authority (#2020060), and Kristian Gerhard Jebsen Stiftelsen (SKGJ-MED-021).

## Author contribution statement

Lars A. R. Ystaas – Conceptualization, Formal Analysis, Data Curation, Visualization, Writing – Original Draft, Writing – Review & Editing.

Pravesh Parekh – Data Curation, Writing – Review & Editing.

Nadine Parker –Writing – Review & Editing.

Ibrahim Akkouh – Writing – Review & Editing.

Viktoria Birkenæs – Writing – Review & Editing.

Ida E. Sønderby – Writing – Review & Editing.

Elise Koch – Data Curation, Writing – Review & Editing.

Espen Hagen – Software, Resources, Writing – Review & Editing.

Oleksandr Frei – Methodology, Writing – Review & Editing.

Alexey Shadrin – Methodology, Formal Analysis, Writing – Review & Editing.

Ole A. Andreassen – Supervision, Funding Acquisition, Project Administration, Writing – Review & Editing.

Kevin S. O’Connell – Conceptualization, Supervision, Funding Acquisition, Writing – Review & Editing.

All authors contributed to the writing of the manuscript and have approved the final version.

