## Supplemental figures for "Genetic liability to addiction underlies comorbid bipolar and substance use disorders"

Lars A. R. Ystaas, Pravesh Parekh, Nadine Parker, Ibrahim Akkouh, Viktoria Birkenæs, Ida E. Sønderby
Elise Koch, Espen Hagen, Oleksandr Frei, Alexey Shadrin, Ole A. Andreassen, Kevin S. O'Connell

### 1 MiXeR

#### 1A Bivariate MiXeR

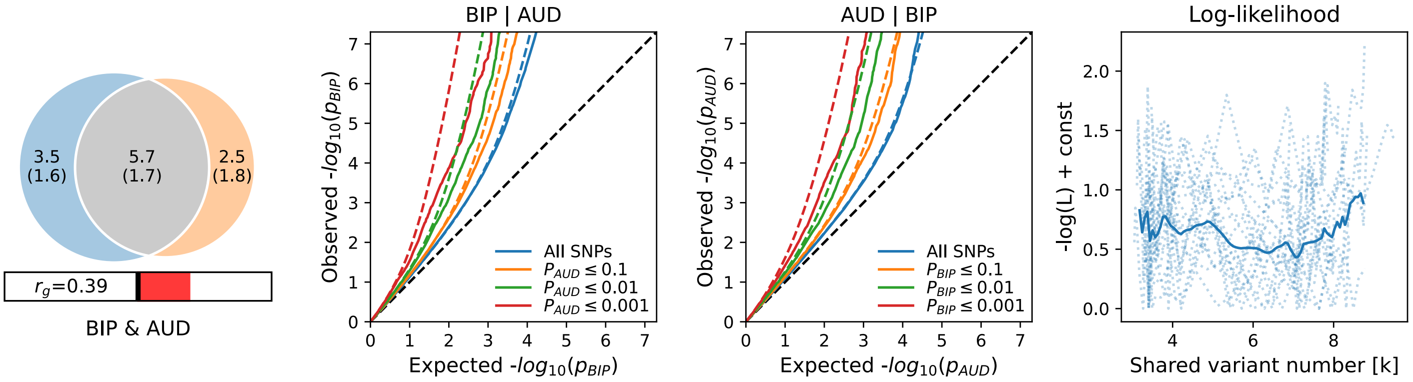

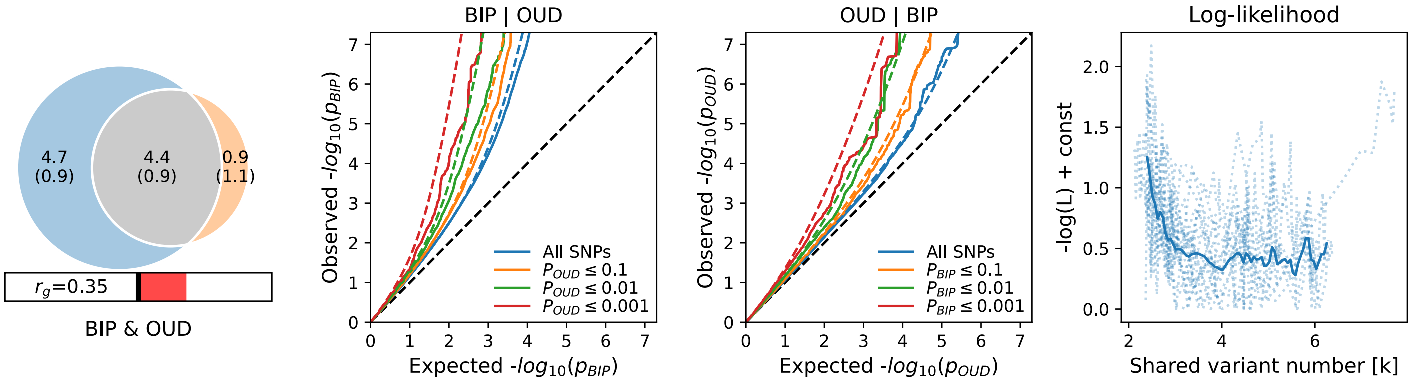

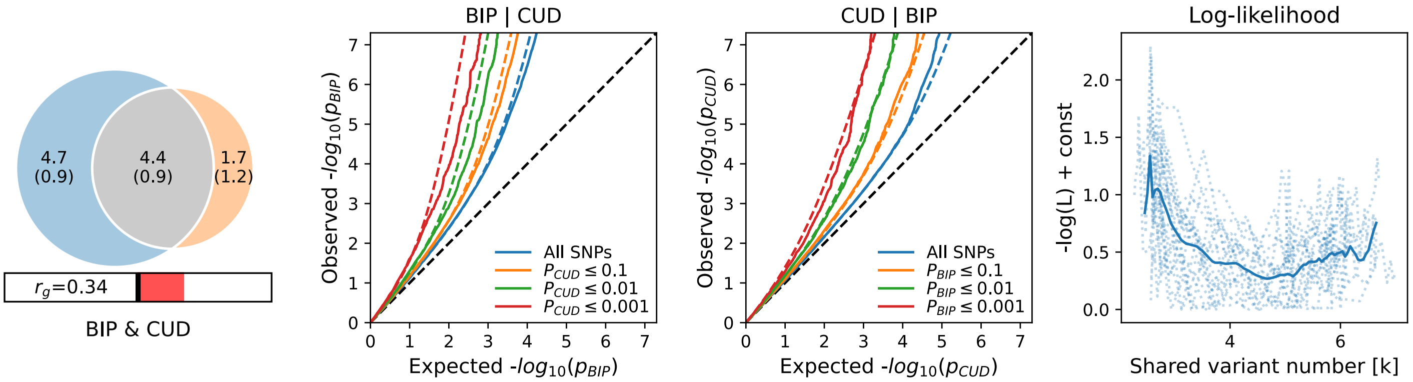

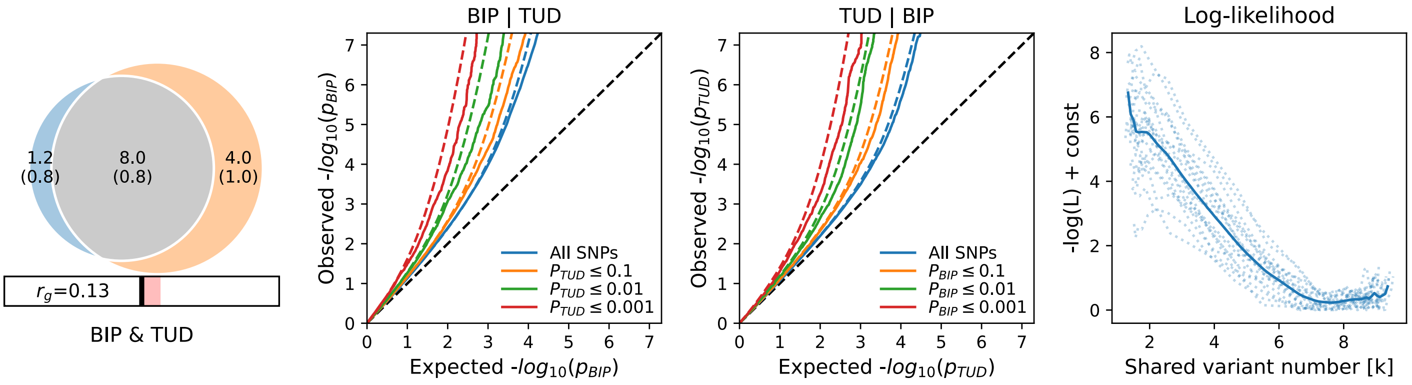

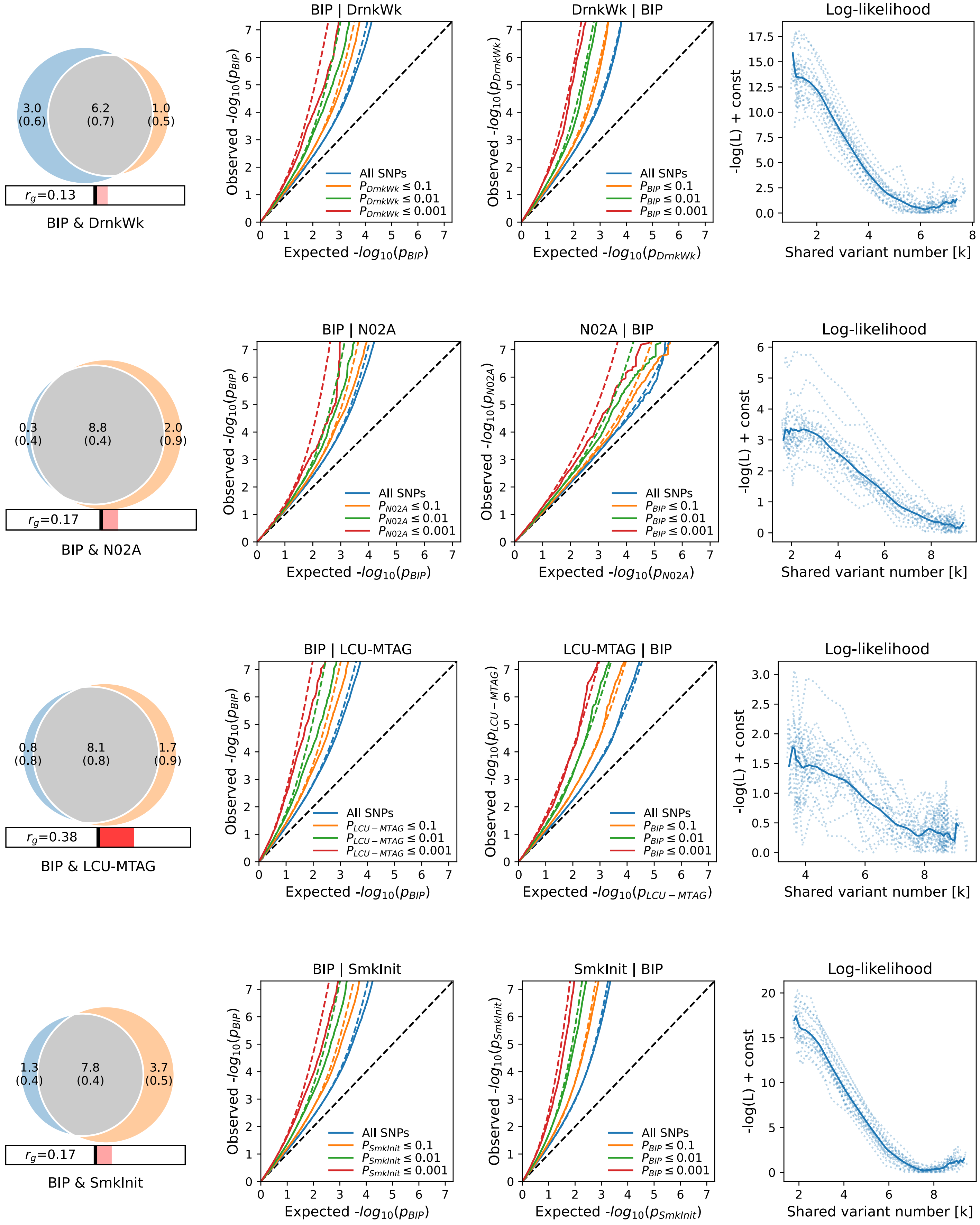

N02A: Prescription opioid use (PrOU).

### 2 GenomicSEM

#### 2A GWAS-by-subtraction: General SU, Unique SUD

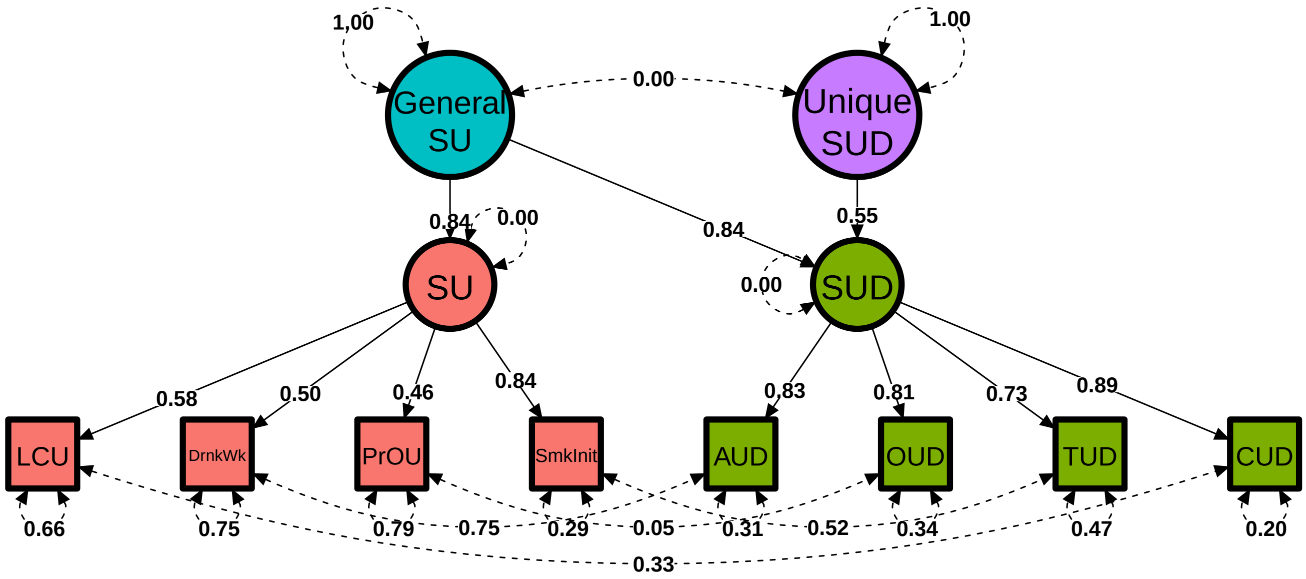

### 3 Polygenic associations in MoBa

#### 3A PRS analyses: Main diagnoses (General SUD, Unique SU)

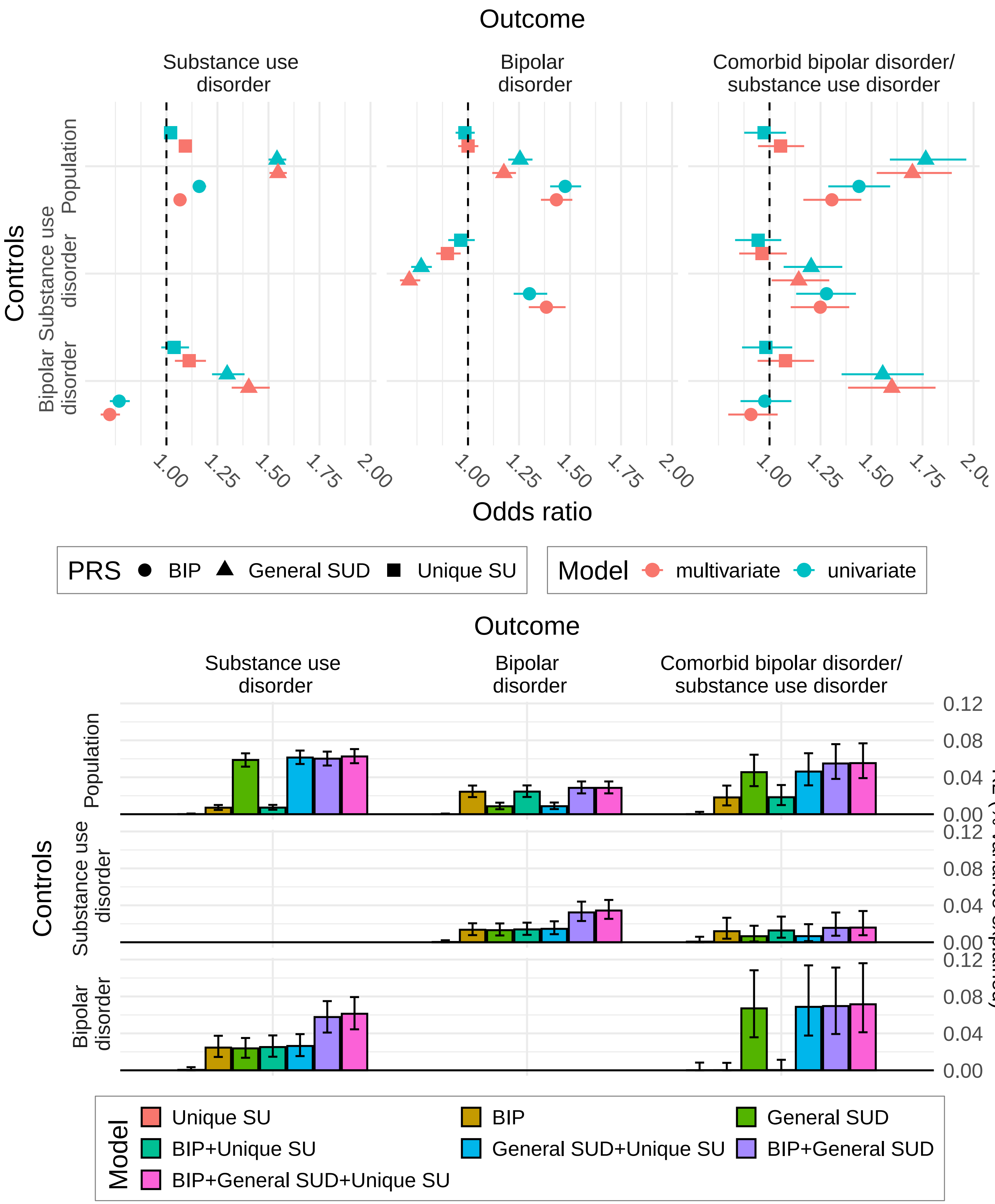

*PRS associations with diagnosis in MoBa, against population, SUD or BIP controls. Error bars denote bootstrap confidence intervals. Odds ratio for 1 SD. increase in PRS value. Univariate models: Single PRS. Multivariate models: All three PRS. All models included covariates as described in Methods.*

#### 3B PRS analyses: Bipolar subgroups (General SUD, Unique SU)

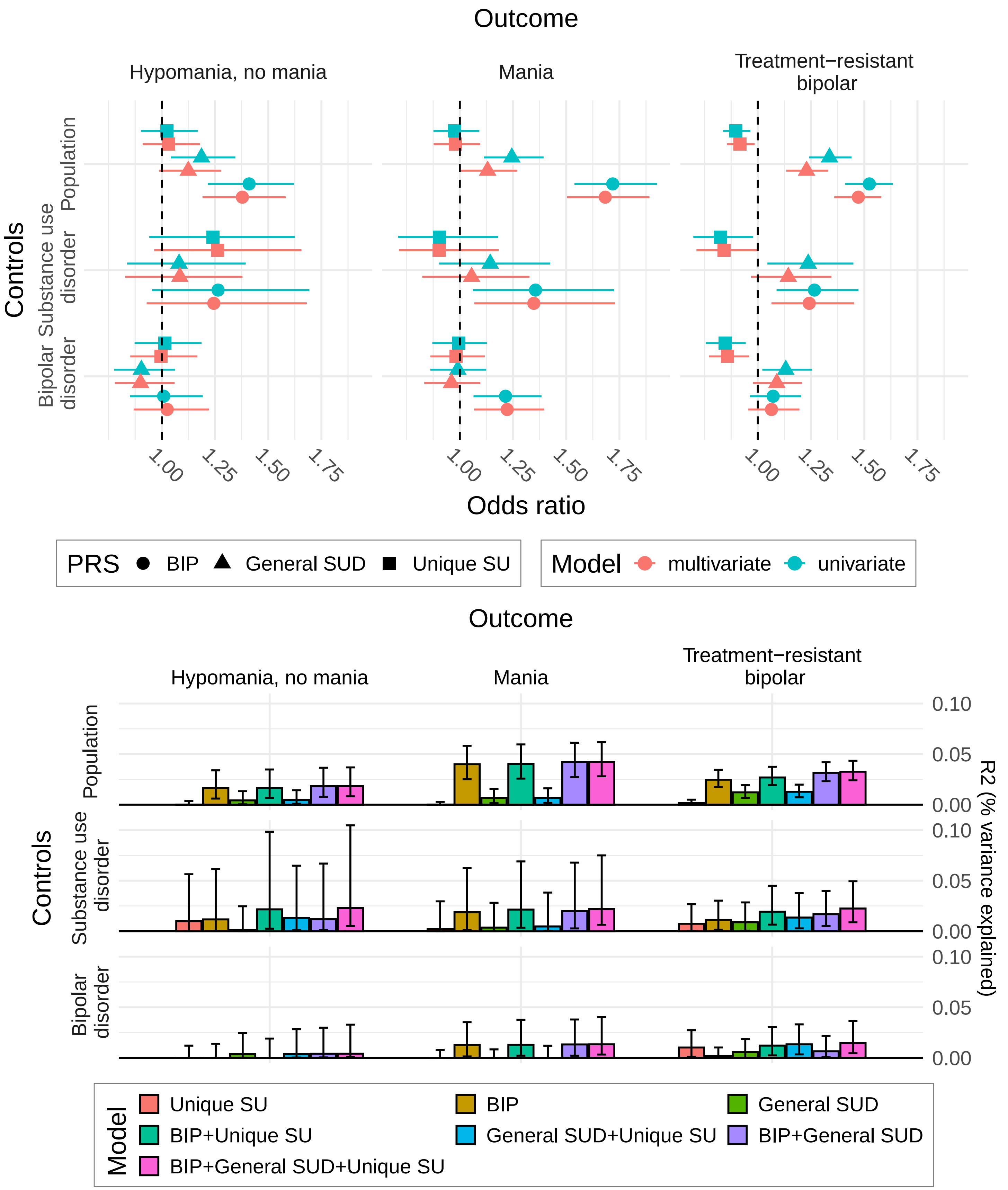

*PRS associations with BIP diagnostic subtypes in MoBa participants. "Hypomania, no mania" refers to individuals with lifetime diagnosis of hypomania, but no mania diagnosis, while "Mania" refers to anyone with a lifetime diagnosis of mania. Error bars denote bootstrap confidence intervals. Odds ratio for 1 SD. increase in PRS value. Univariate models: Single PRS. Multivariate models: All three PRS. All models included covariates as described in Methods.*

##### 3C PRS analyses: Self-reported SU (General SUD, Unique SU)

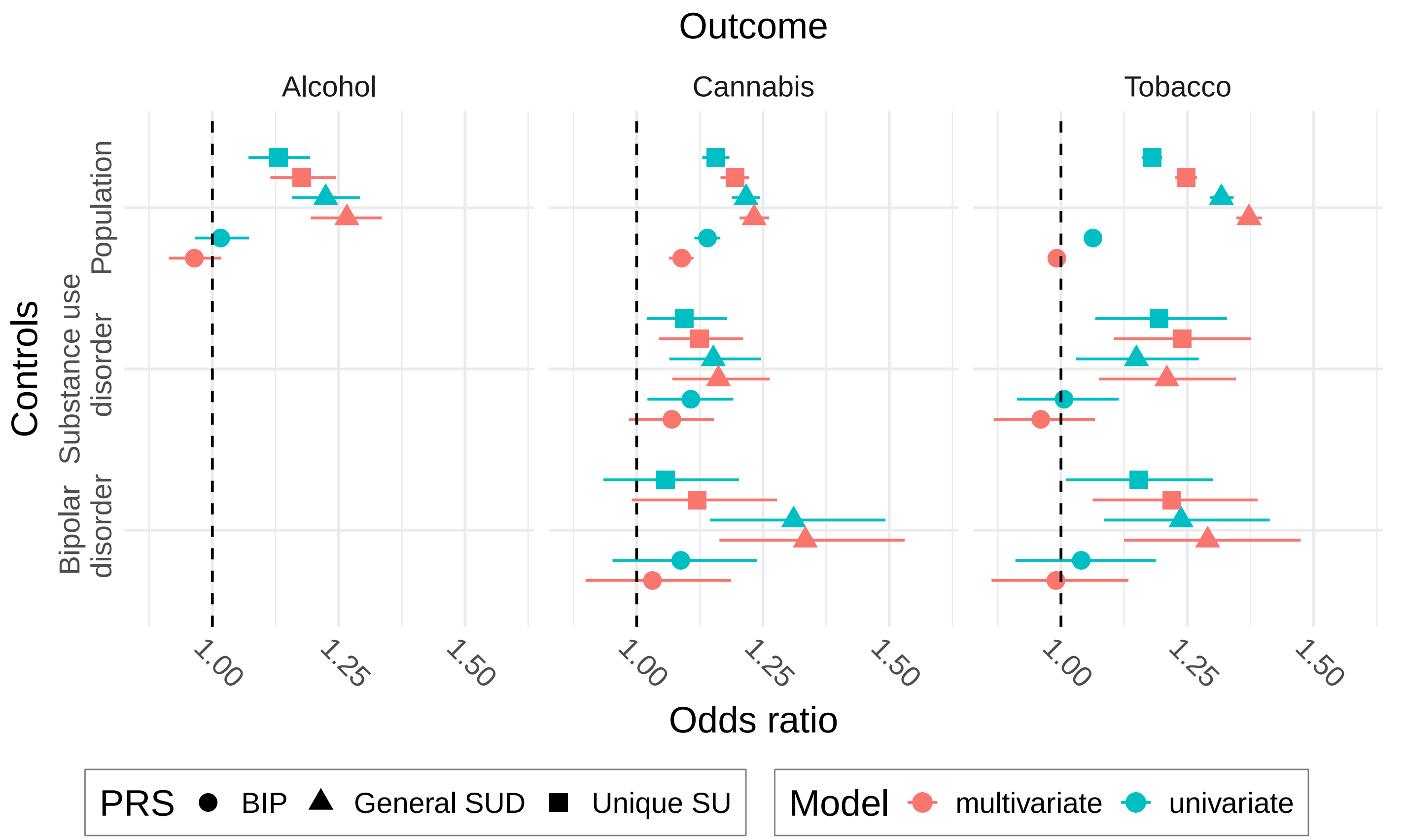

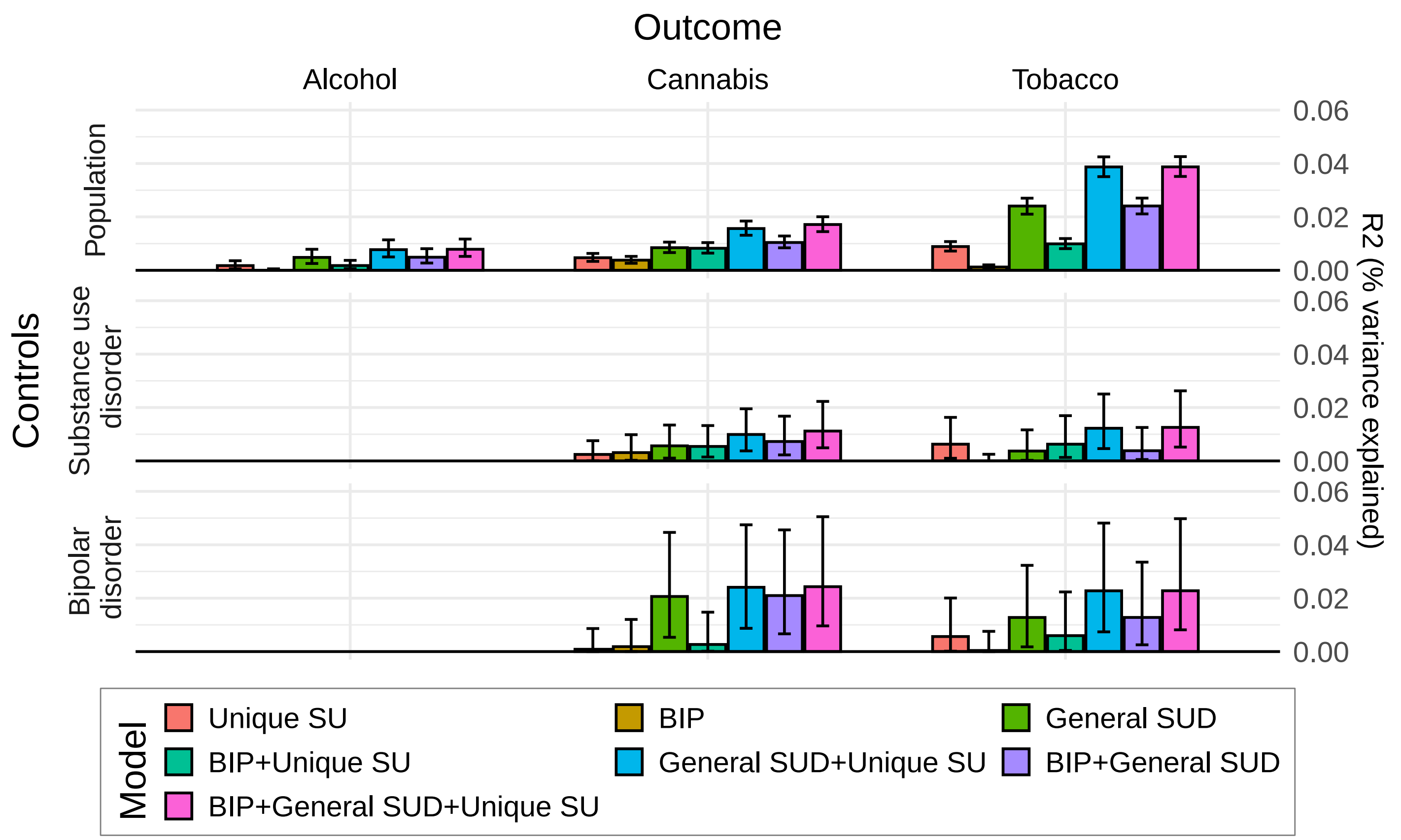

*PRS associations with self-reported lifetime substance use in MoBa adults within general population, BIP and SUD cases. Error bars denote bootstrap confidence intervals. Due to small numbers of individuals in the dataset without lifetime alcohol use, models in subgroups for this phenotype was not able to be estimated. Odds ratio for 1 SD. increase in PRS value. Univariate models: Single PRS. Multivariate models: All three PRS. All models included covariates as described in Methods.*

##### 3D PRS analyses: Self-reported SU adolescents (General SUD, Unique SU)

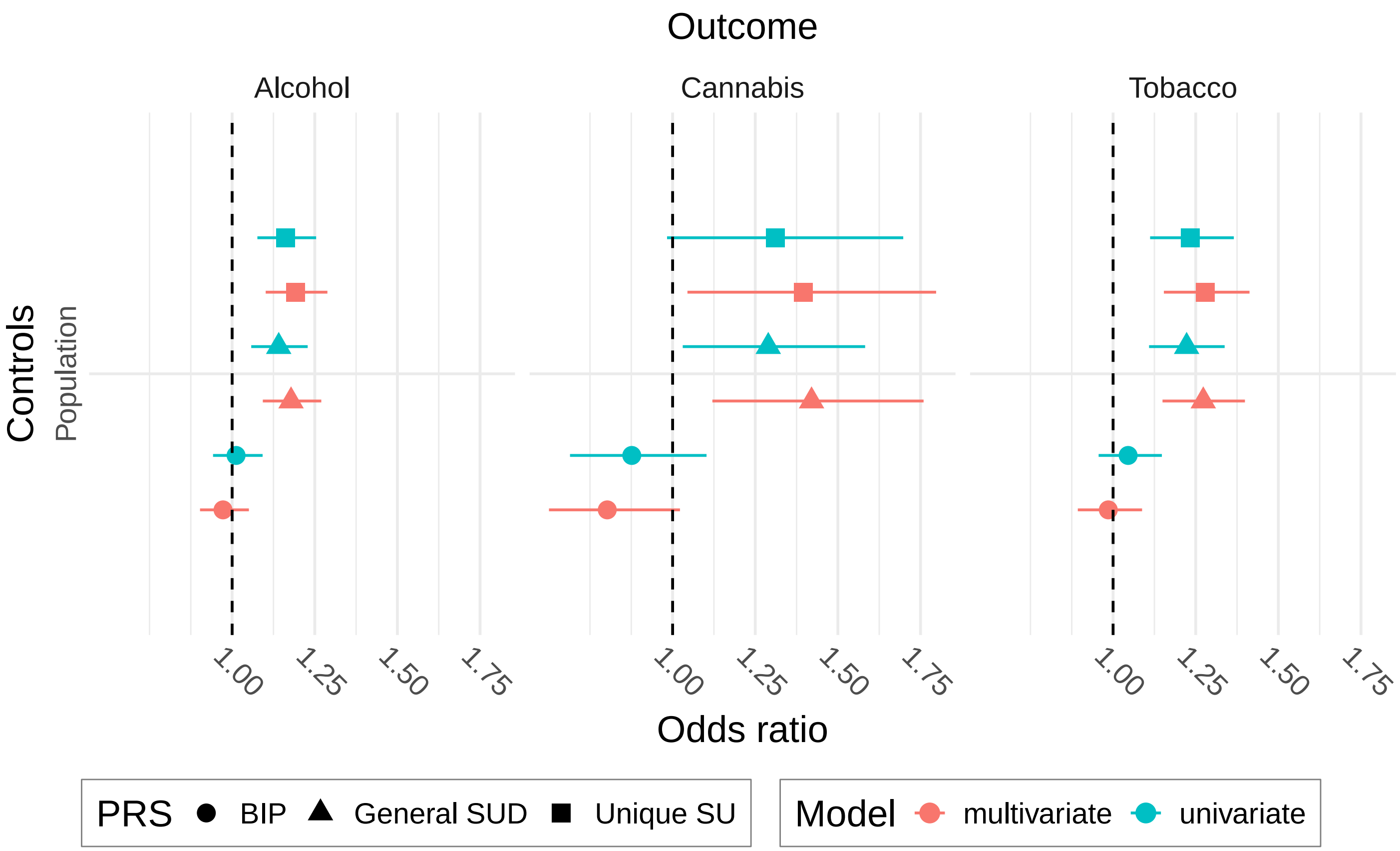

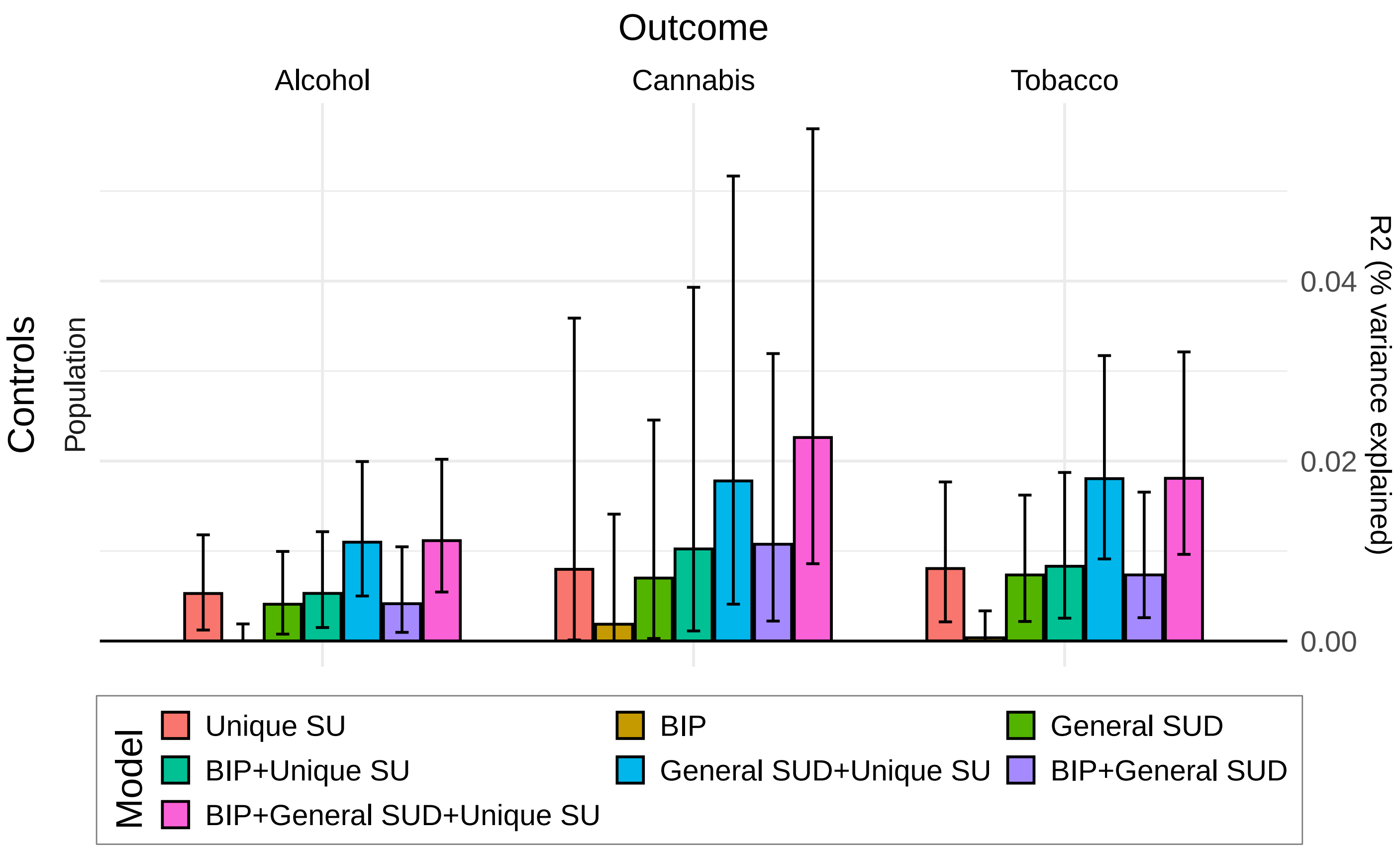

*PRS associations with self-reported lifetime substance use in MoBa adolescents within general population, BIP and SUD cases. Error bars denote bootstrap confidence intervals. Due to small numbers of adolescents with diagnosed BIP and SUD, models in subgroups for these phenotypes were not able to be estimated. Odds ratio for 1 SD. increase in PRS value. Univariate models: Single PRS. Multivariate models: All three PRS. All models included covariates as described in Methods.*

##### 3E PRS analyses: Main diagnoses (General SU, Unique SUD)

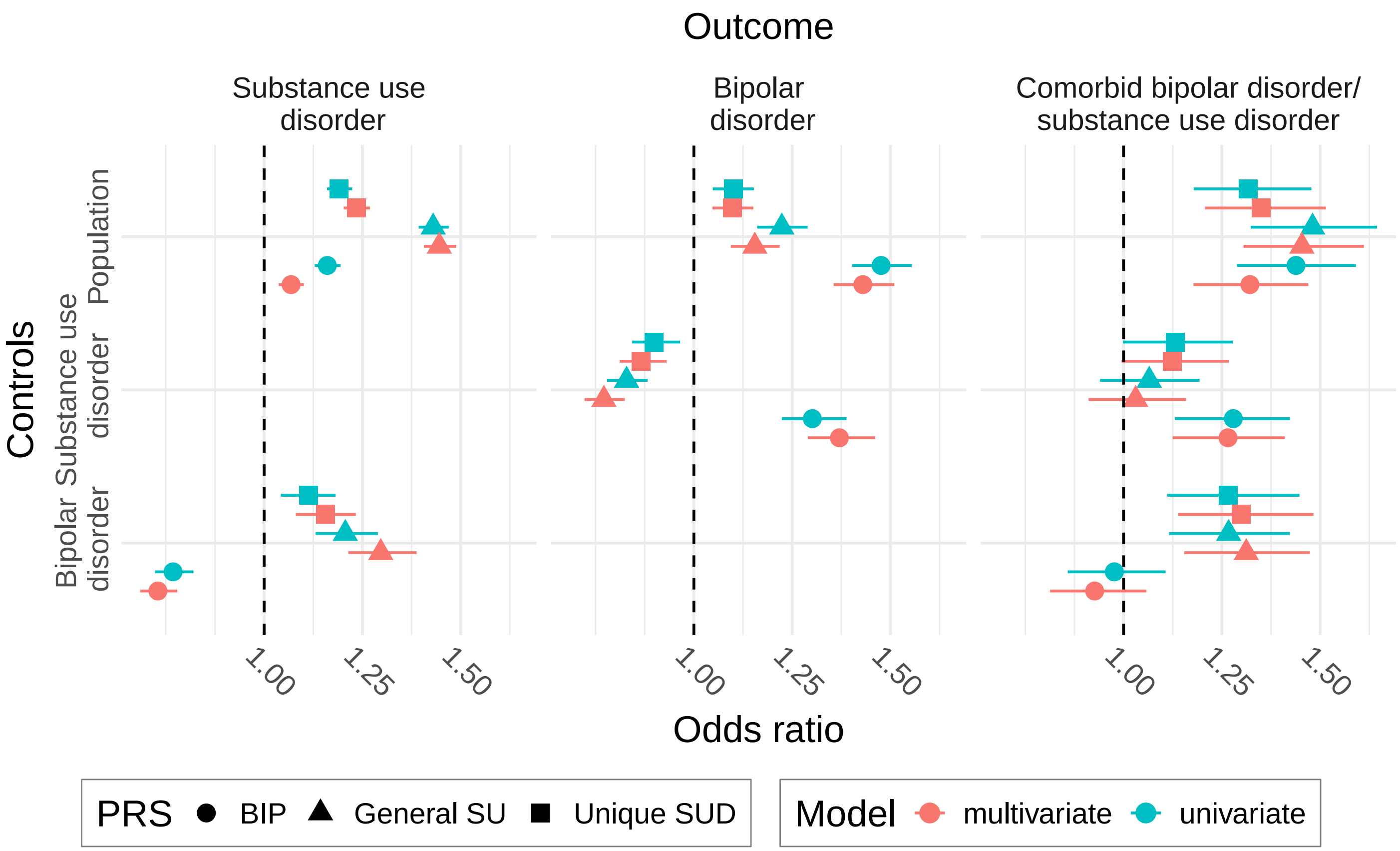

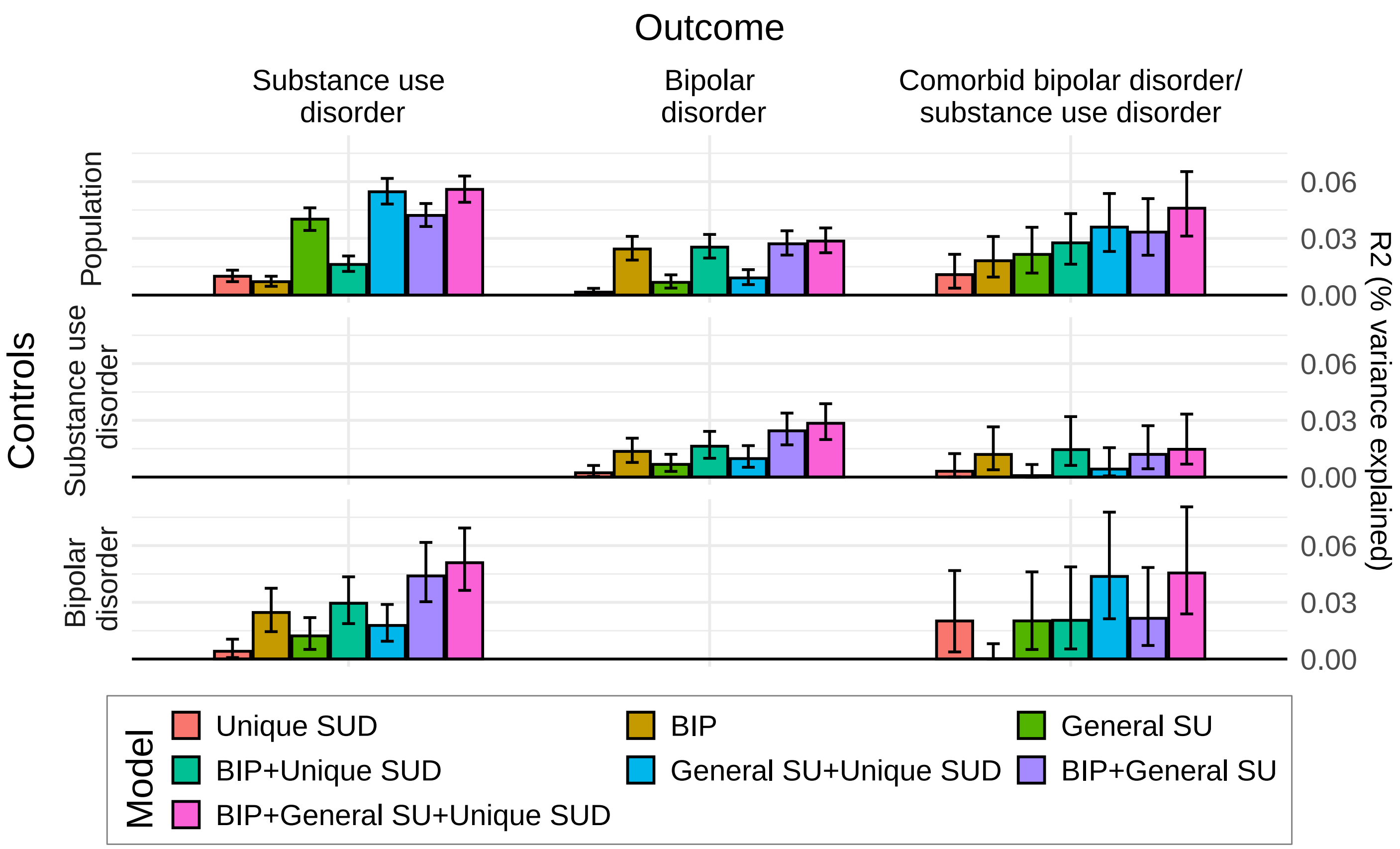

*PRS associations with diagnosis in MoBa, against population, SUD or BIP controls. Error bars denote bootstrap confidence intervals. Odds ratio for 1 SD. increase in PRS value. Univariate models: Single PRS. Multivariate models: All three PRS. All models included covariates as described in Methods.*

#### 3F PRS analyses: Bipolar subgroups (General SU, Unique SUD)

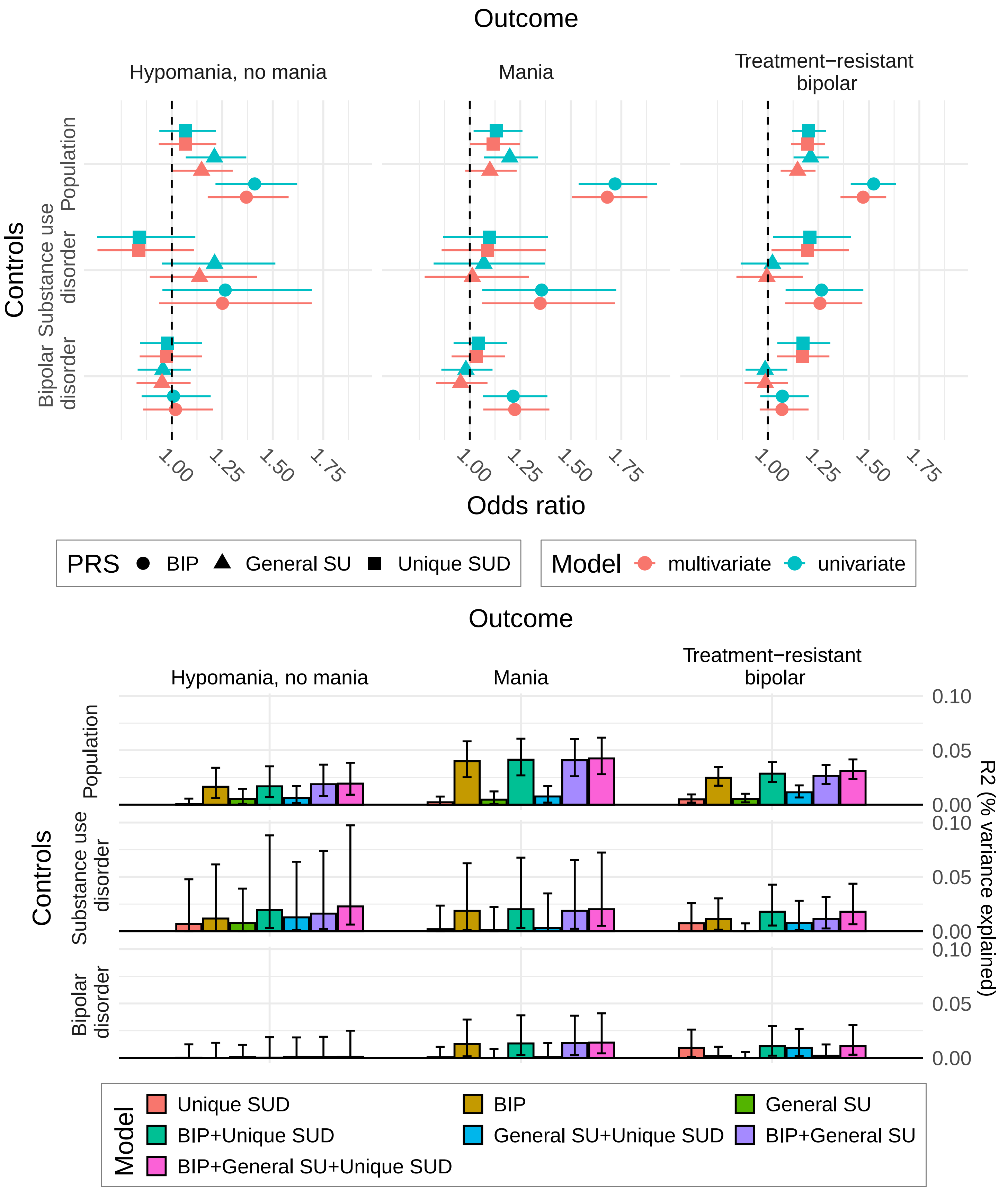

*PRS associations with BIP diagnostic subtypes in MoBa participants. "Hypomania, no mania" refers to individuals with lifetime diagnosis of hypomania, but no mania diagnosis, while "Mania" refers to anyone with a lifetime diagnosis of mania. Error bars denote bootstrap confidence intervals. Odds ratio for 1 SD. increase in PRS value. Univariate models: Single PRS. Multivariate models: All three PRS. All models included covariates as described in Methods.*

##### 3G PRS analyses: Self-reported SU adults (General SU, Unique SUD)

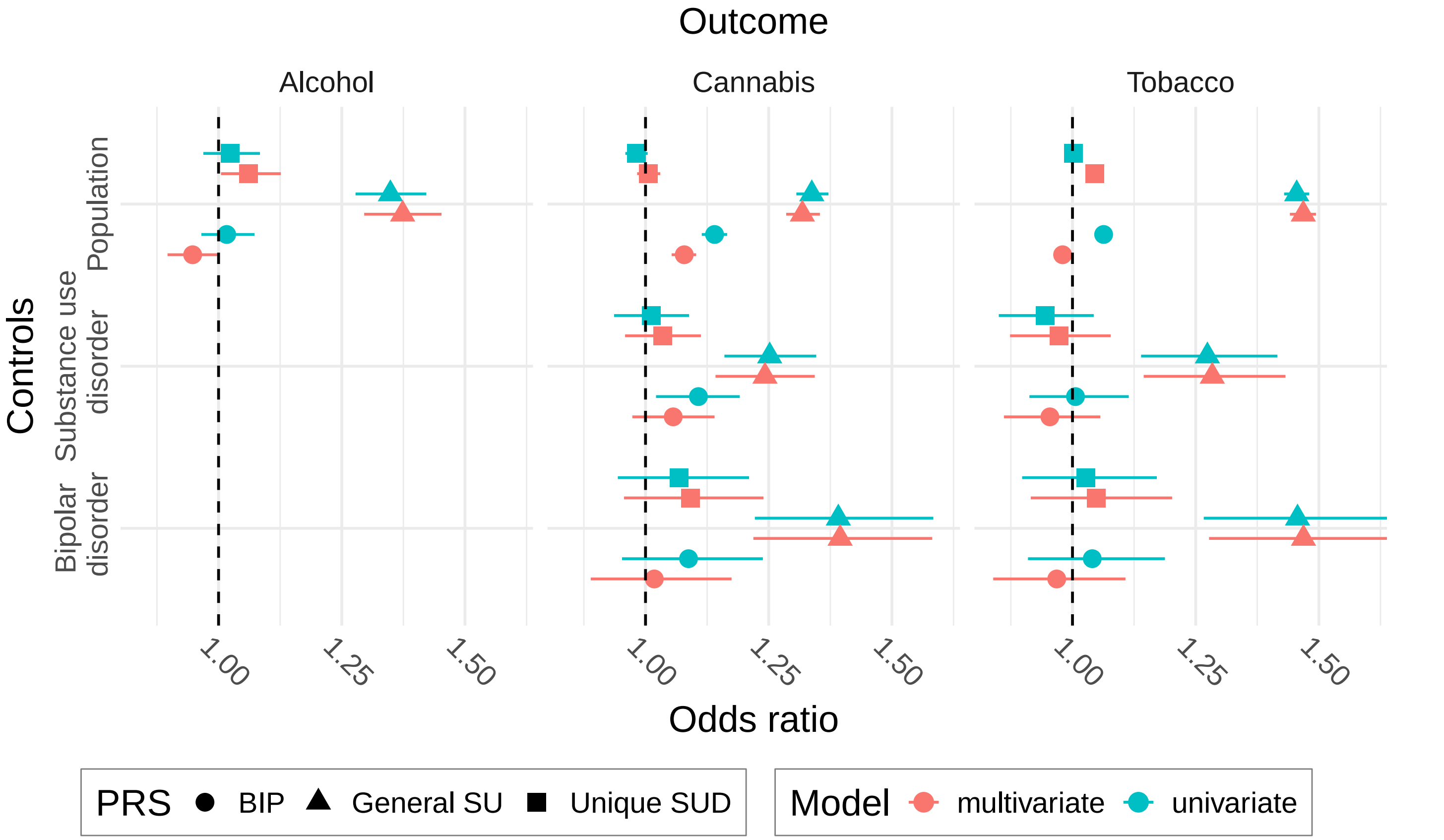

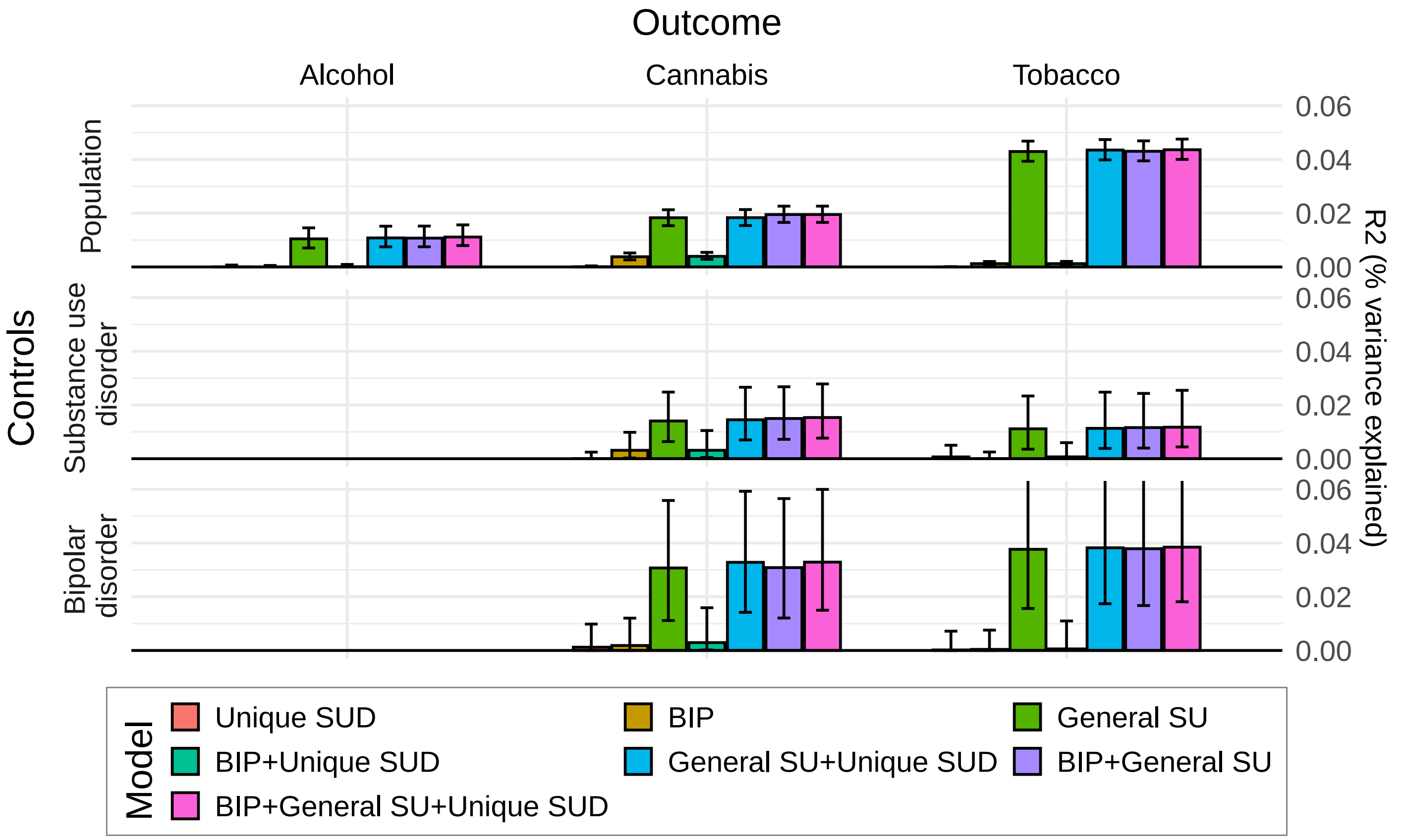

*PRS associations with self-reported lifetime substance use in MoBa adults within general population, BIP and SUD cases. Error bars denote bootstrap confidence intervals. Due to small numbers of individuals in the dataset without lifetime alcohol use, models in subgroups for this phenotype was not able to be estimated. Odds ratio for 1 SD. increase in PRS value. Univariate models: Single PRS. Multivariate models: All three PRS. All models included covariates as described in Methods.*

##### 3H PRS analyses: Self-reported SU adolescents (General SU, Unique SUD)

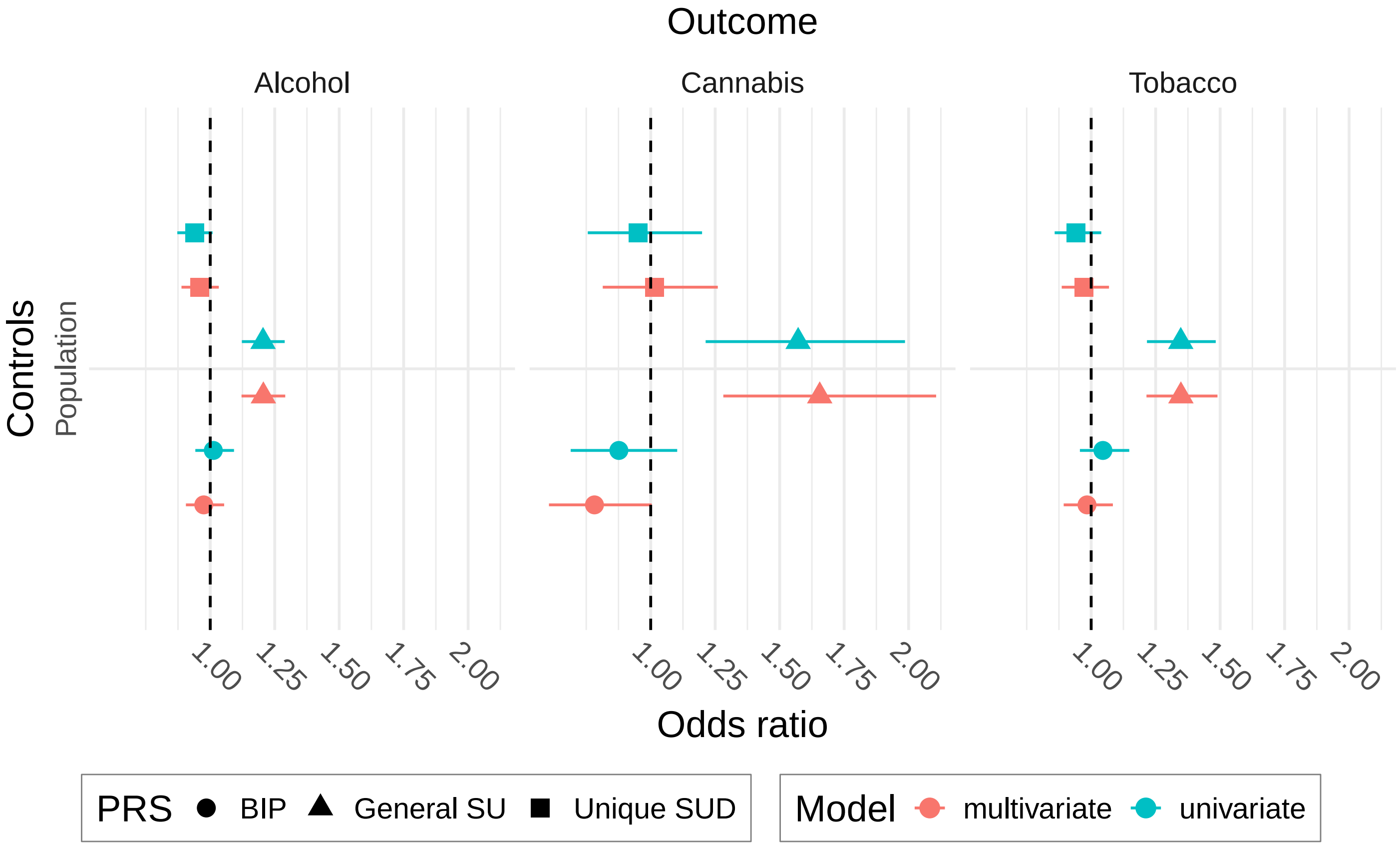

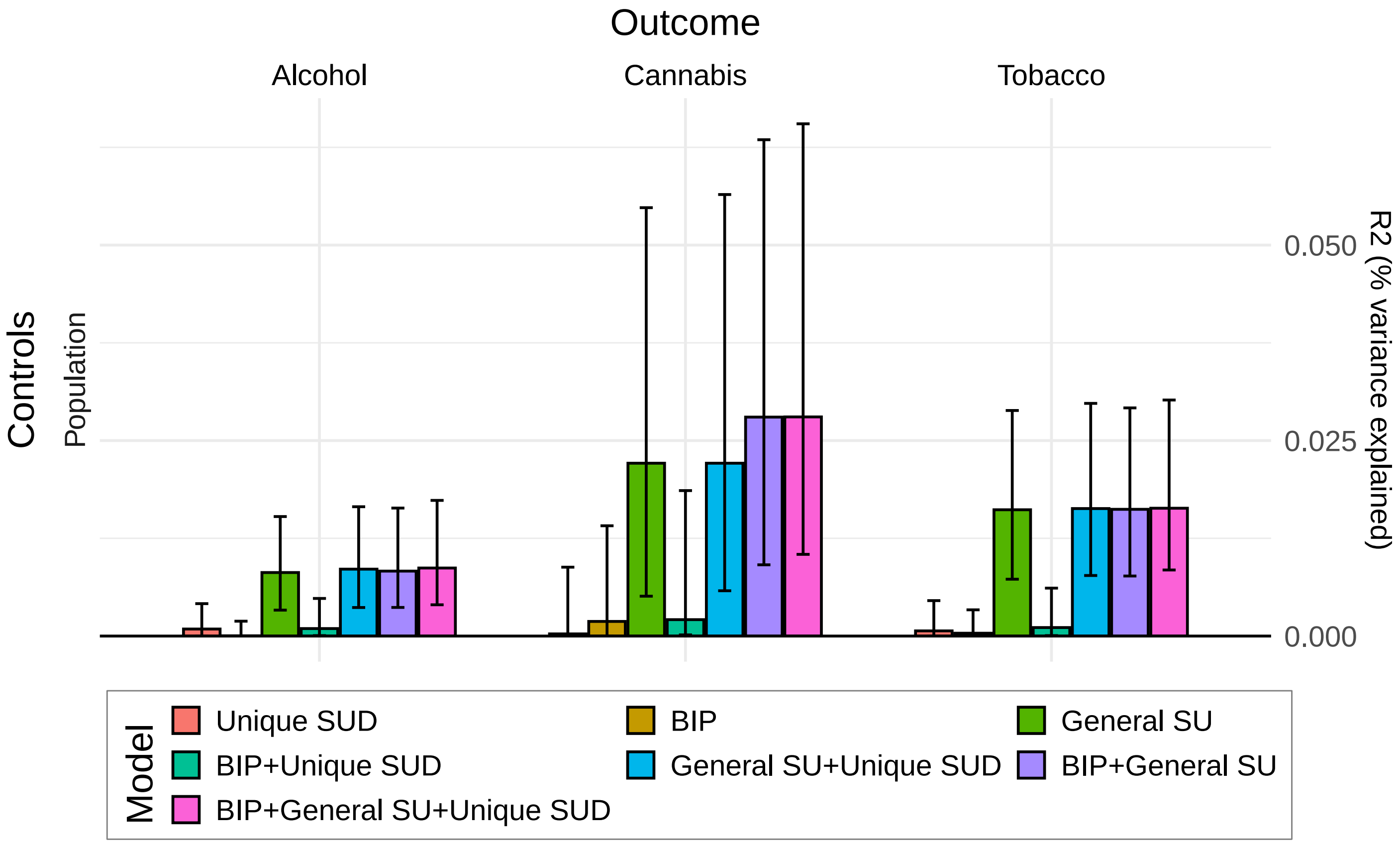

*PRS associations with self-reported lifetime substance use in MoBa adolescents within general population, BIP and SUD cases. Error bars denote bootstrap confidence intervals. Due to small numbers of adolescents with diagnosed BIP and SUD, models in subgroups for these phenotypes were not able to be estimated. Odds ratio for 1 SD. increase in PRS value. Univariate models: Single PRS. Multivariate models: All three PRS. All models included covariates as described in Methods.*

#### 3I PRS distributions

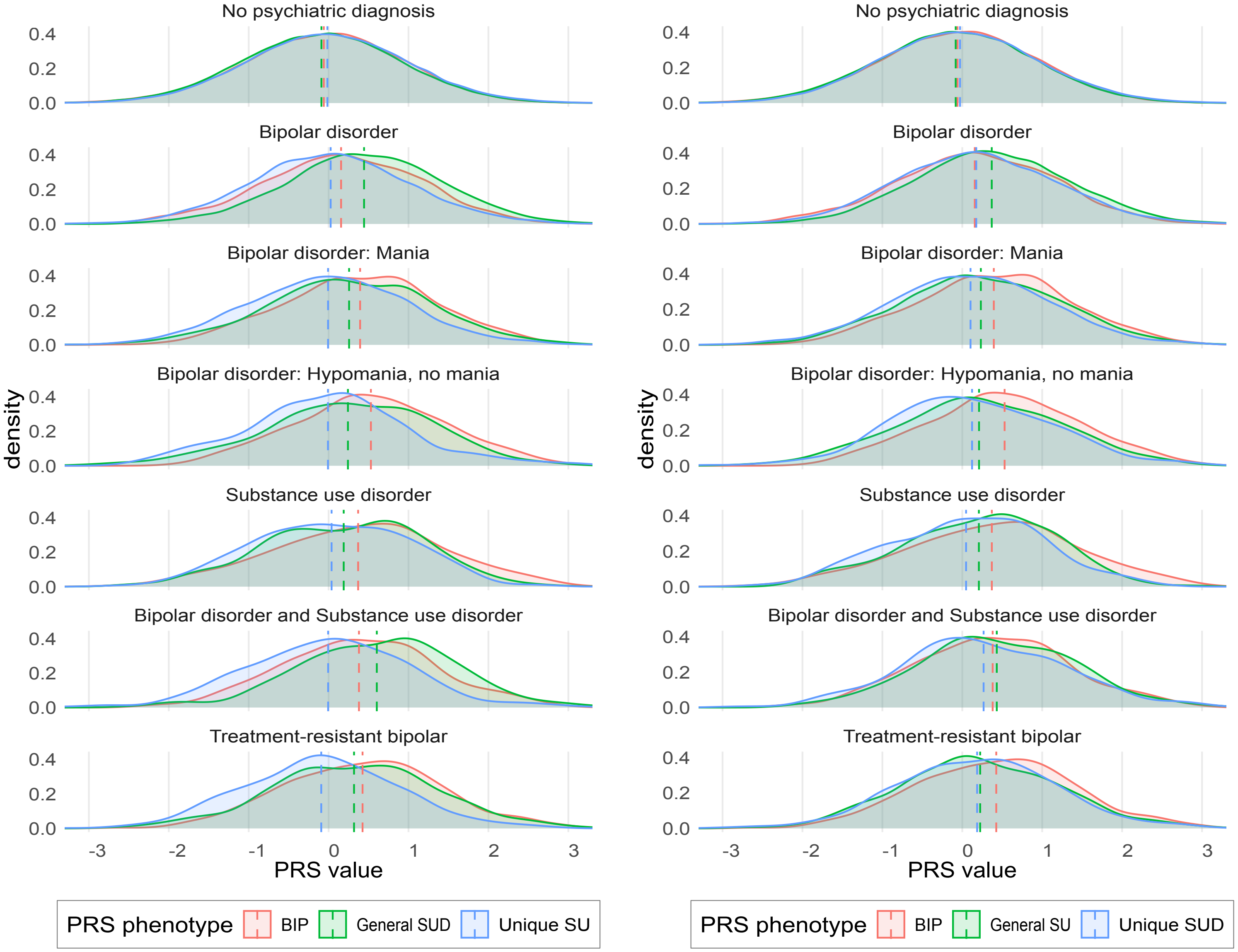

*Smoothed distributions of PRS values in diagnostic groups in MoBa. Dotted vertical lines denote mean value.*
