## Supplemental methods for "Genetic liability to addiction underlies comorbid bipolar and substance use disorders"

Supplementary methods

Lars A. R. Ystaas^1,2^, Pravesh Parekh^1,5^, Nadine Parker^1,2^, Ibrahim Akkouh^1,4^, Viktoria Birkenæs^1^, Ida E. Sønderby^1,,4^, Elise Koch^1,7^, Espen Hagen^1^, Oleksandr Frei^1,2,6^, Alexey Shadrin^1,3^, Ole A. Andreassen^1,2,3,4^, Kevin S. O’Connell^1,2^

Affiliations:

1. Centre for Precision Psychiatry, Institute of Clinical Medicine, University of Oslo
2. Division of Mental Health and Addiction, Oslo University Hospital, Oslo, Norway
3. KG Jebsen Centre for Neurodevelopmental Disorders, University of Oslo, Oslo, Norway.
4. Department of Medical Genetics, Oslo University Hospital, Oslo, Norway
5. Center for Multimodal Imaging and Genetics, J. Craig Venter Institute, La Jolla, CA, USA
6. Center for Bioinformatics, Department of Informatics, University of Oslo, Oslo, Norway
7. Center for Psychopharmacology, Diakonhjemmet Hospital, Oslo, Norway

GenomicSEM

GenomicSEM estimates a SNP-level heterogeneity statistic, Q_SNP_, with larger values indicating that the SNP effects of a factor are not mediated by the factor itself, but are driven by a subset of the indicator traits included in the factor(1). To remove heterogeneous SNPs, the variants were clumped on Q_SNP_ p-value using PLINK(2). To be considered heterogeneous, the SNPs had to be in LD (r^2^ > 0.6) with significantly heterogeneous SNPs (p(Q_SNP_) < 5e-8), using a 250kb window. These heterogeneous SNPs were removed from downstream analyses to ensure effects were related to the latent factor and not specific input traits.

Questionnaires and registry data

Questionnaires are sent out to the parents and the child at specific time points of the child and the parent’s life. We have focused on the first questionnaire filled out by the parents in week 15 of the pregnancy and the questionnaire completed when the child is 14 years old as this is the first time there are items related to substance use for the offspring. Questions relating to substance use are answered by the child. The current study is based on version 12 of the quality-assured data files released for research in 2019.

We obtained diagnostic data from the Norwegian Patient Registry (NPR), Control and Payment of Health Reimbursements Database (KUHR) and Municipal Patient and User Register (KPR) which together provide comprehensive health records for the Norwegian population. These registries cover consultations across public hospitals, outpatient facilities, and contract specialists in Norway, in effect ensuring nearly complete coverage of patient treatment in the country within the specified time we have available data – in our case between the periods 2006-October 2023 (KUHR), July 2016-2024 (KPR) and 2008-2024 (NPR). The registries include detailed medical diagnoses coded according to the International Classification of Diseases (ICD-10)(3). KUHR and KPR also include general practitioner diagnoses in the ICPC-2 diagnostic system(4). Prescription data was gathered from the Norwegian Prescribed Drug Registry (LMR), with data covering all outpatients who have collected a prescription from a Norwegian pharmacy from 2004 until 2024.
